# Regime type and Data Manipulation: Evidence from the COVID-19 Pandemic

**DOI:** 10.1101/2022.12.11.22283310

**Authors:** Simon Wigley

## Abstract

Autocratic and democratic leaders have an incentive to misreport data that may reveal policy failure. However, it is easier for autocratic leaders to fabricate data because they are not subject to scrutiny from media, opposition parties, and civil society. This suggests that autocratic governments are more likely to manipulate policy-relevant statistics than democratic governments. It is inherently difficult to test that claim because researchers typically do not have access to data from sources other than the government. The COVID-19 pandemic represents a unique opportunity to examine the relationship between regime type and data manipulation because of its widespread impact, as well as the ability to compare reported with excess deaths and test for statistical anomalies in reported data. Based on regressions for undercounting and statistical irregularities that take into account unintentional mismeasurement, I find that autocratic governments are more likely to deliberately under-report the impact of COVID-19 than their democratic counterparts.

## Introduction

Are autocratic governments more likely to manipulate policy-relevant data than democratic governments? According to one view, autocratic and democratic leaders both have an incentive to misreport data when the truth may reveal incompetence, leading to protest or electoral losses. However, data manipulation is harder to achieve in a democracy because the government is subject to greater scrutiny from opposition parties, independent media, and civil society organizations (Carlitz & McLellan, 2021; Hollyer et al., 2011). According to another view, autocratic governments are primarily concerned about the interpretation of information, rather than access to information (Rozenas & Stukal, 2019). Autocratic leaders may calculate that bad news will not lead to collective action if they can persuade citizens that the government was not responsible. Autocrats can, for example, use their control over media and the internet to convince citizens that a bad outcome is due to external forces (e.g., global macroeconomic factors) or natural phenomena (e.g., pandemic) that are beyond the government’s control.

It is not immediately obvious, therefore, that autocratic leaders are more likely to provide inaccurate data than their democratic counterparts. In this study I argue that autocrats retain an incentive to manipulate data even as they seek to shift the blame to external forces or natural phenomena. This is because they cannot be sure whether their attempts to persuade citizens that they are not responsible will succeed. They have a greater capacity to shape perceptions than democratic leaders, but there is a risk that a critical number of citizens remain unconvinced, leading to criticism and protest. Democratic leaders, by contrast, are less able to successfully deploy either strategy - manipulate data or shift the blame - because they lack control over traditional and online media, opposing political parties, and civil society groups.

It is inherently difficult to prove whether autocrats are more likely to manipulate because data are typically not available from a source that is independent of the government. Historically, national statistical agencies emerged in order to serve the aims of government (Brambor et al., 2020; Brewer, 1990, Chapter 8), and they often remain subject to political pressure or direct control (Herrera & Kapur, 2017, pp. 375–376). Various methods have been developed to enable researchers to detect manipulation when government-independent data is not available. Firstly, they may use a substitute indicator to estimate the real values (e.g., night time lights as a proxy for GDP) (Magee & Doces, 2015; Martínez, 2022). However, that strategy can only be used in those cases where a suitable proxy is available. Secondly, they may check the reported data for statistical anomalies. For example, they may examine whether the digits are consistent with Benford’s Law (Michalski & Stoltz, 2013), or whether a count sequence is unexpectedly smooth (Kobak, 2022). However, attentive autocrats can manipulate the data such that they accord with the expected pattern. Thirdly, researchers may infer from aggregate level data that has been provided by the government. For example, they may estimate excess deaths due to a shock or policy intervention by comparing total mortality from all causes with past trends (Ashton et al., 1984; Spagat & van Weezel, 2017). However, that method is dependent on the availability of complete and reliable data at the aggregate-level. Fourthly, they may estimate the real data values for a particular country using statistical models that reliably predict the outcome of interest in those countries where data is known to be complete and accurate (Viboud et al., 2016).This method is reliant on the availability of veridical data for the variables that are included in the predictive model. Governments may be willing to provide accurate measures of those variables if, by themselves, they do not reveal incompetence. Alternatively, data for those predictor variables may be available from sources independent of the government.

While all of these methods have their shortcomings, each enables the researcher to get closer to the truth, especially in those cases where tampering is suspected. Each provides a way to reduce the likelihood that they are drawing incorrect conclusions about the phenomena under study. In the analysis below, I rely on data that have been constructed based on a combination of the last two methods (aggregate trends and predictive model), as well as the second method (testing for statistical anomalies).

The devastating pandemic produced by the spread of severe acute respiratory syndrome coronavirus 2 (SARS-CoV-2) represents a unique opportunity to examine whether autocratic governments are more susceptible to data manipulation than their democratic counterparts. Firstly, the sheer scale of the shock and, therefore, the potential threat posed to the legitimacy of each government in the eyes of their citizens, magnifies the incentive for political leaders to hide the truth. Secondly, excess mortality can be estimated for most sovereign states based on a combination of predictive models and a comparison between total mortality trends before and after the onset of the pandemic. The deviation of reported COVID-19 deaths from excess deaths provides an estimate of the extent to which governments are, deliberately or unintentionally, undercounting. If we control for the factors that have led to unintentional undercounting, then we can estimate the degree of manipulation and whether it varies by regime type. Furthermore, we can directly test for manipulation by checking whether there are statistical anomalies in the daily reported cases and deaths. Deviation of those reported numbers from Benford’s Law or expected variation across time allows us to detect manipulation and assess whether it varies by regime type. Thirdly, the pandemic has impacted virtually every country in the world and so we can examine the phenomenon of data manipulation for a large sample of countries. In the following analysis I utilize excess mortality data for as many as 197 states and statistical anomaly estimates for as many as 201 states.

Previous studies on the impact of regime type on data manipulation have mostly focused on the reporting of data in autocratic contexts (e.g. Carlitz & McLellan, 2021; Chen et al., 2019; Lamberova & Sonin, 2022; Wallace, 2022, Chapter 6). They do not, therefore, provide a systematic comparison between data manipulation in autocratic and democratic countries. Hollyer et al (2011) represents a partial exception, but their focus is on the withholding of policy-relevant data, rather than the misreporting of such data. As Carlitz and McLellen (2021) note, many autocracies are now more willing to provide development data due to the expectations of foreign aid donors, as well as the targets specified in the Millennium Development Goals and Sustainable Development Goals. The question is whether they accurately report such data.

Magee and Docees (2015) and Martínez (2022) investigate the impact of regime type on the manipulation of economic data for a large number of democratic and autocratic countries. Using night time lights as a proxy for GDP, both studies find that autocratic regimes tend to overstate yearly GDP growth. In the current study I shift the focus to the reporting of population health statistics.^1^ Three recent studies examine the relationship between regime type and the manipulation of COVID-19 data using Benford’s law (Kilani, 2021) and the discrepancy between reported and excess mortality (Knutsen & Kolvani, 2022; Neumayer & Plümper, 2022) for a large sample of countries. In this study I build on those earlier results by using three distinct measures of manipulation (ratio of excess to reported deaths, Benford-noncompliance of daily counts, and underdispersion of daily counts). Moreover, for the sake of further robustness, I use four different estimates of excess mortality, as well as five different measures of democracy. This multi-measurement approach helps to reduce the likelihood that the overall conclusions of this study an artifact of a particular estimation model. Finally, I examine whether any of core components of democratic rule – free and fair elections, freedom of association, freedom of expression, suffrage, and elected officials - have an outsized influence on the level of data manipulation.

In the following, I firstly elaborate on the theory behind the claim that autocratic governments manipulate policy-relevant statistics more than democratic governments. I then describe those factors independent of regime type that increase the likelihood that COVID-19 deaths will go under-reported. I then describe the estimation models, results, and 12 robustness checks. In the penultimate section I discuss how the results align with the existing literature on the link between regime survival and information control, as well as the limitations of this study. I conclude by outlining the problems that data manipulation creates for citizens, researchers, and international organizations, as well as potential solutions.

### Why do autocratic leaders manipulate reported data more than democratic leaders?

Autocratic leaders have an incentive not to share accurate information if it might lead to their removal via mass mobilization. That is, full and accurate disclosure may enable citizens, firstly, to become aware of any failure in government policy and, secondly, to realize that their fellow citizens are also aware of that failure. As a result, each citizen has enough information to judge whether protests will generate sufficient participation to remove the government (Hollyer et al., 2015). Incumbent democratic leaders also have an incentive to tamper with published data when the actual data might threaten their performance at the ballot box. However, it is harder for them to hide poor performance in this way because they are subject to greater scrutiny from opposition parties, civil society, and media. There is a greater likelihood that manipulation by a democratic government will be detected, simultaneously drawing further attention to the negative news it was attempting to hide and tainting its credibility in the eyes of voters.

Autocratic leaders may also attempt to use their control over traditional and online media to persuade citizens that they are not responsible for bad news (Rozenas & Stukal, 2019). ^2^ They may, for example, promote the view that the COVID-19 pandemic is an unstoppable natural phenomenon, or that its spread is due to the containment failures of other countries. Indeed, this may be the preferred approach in those cases where the disclosure of full and accurate data helps to combat the problem. Hiding the severity of an epidemic, for example, may lure citizens into a false sense of security, frustrating attempts to encourage life-saving changes in behavior (e.g., physical distancing, mask-wearing, and vaccination).

However, autocratic leaders cannot solely rely on that strategy because of the risk that it will fail. They have a greater capacity to shape the perceptions of citizens than democratic leaders, but they cannot rule out the possibility that a critical number of citizens will remain unpersuaded, thereby exposing them to criticism and protest. Ironically the lack of communication openness that is needed by autocrats to prevent collective action also makes it harder for them to gauge the proportion of citizens who do not believe their spin (Schedler, 2013, pp. 37–39). Fear of reprisal means the outward behavior or expressed opinion of citizens in an autocratic context may not track whether they actually believe the government’s narrative (Jiang & Yang, 2016; Wedeen, 2015). Under these conditions of uncertainty, autocrats are more likely to conclude that a combination of data manipulation and shifting the blame (i.e. hiding the true extent of the bad news and spinning that news) represents the optimal way to prevent collective action.

Generally, the exact level of data manipulation (and concomitant level of spin) will depend on context. Autocrats may reduce the level of manipulation when citizens can reliably infer the real values (e.g. comparing official inflation statistics with supermarket prices),^3^ or when it starts to threaten the implementation of policy. Too much under-reporting of COVID-19 infections and deaths, for example, may frustrate attempts to encourage physical distancing, mask-wearing and vaccination. In such cases they are more reliant on persuading citizens that they are not responsible. By the same token, they may increase the level of manipulation when citizens find it harder to verify data (e.g. epidemic, conflict, or famine deaths), or it is less likely to hinder the implementation of policy (e.g. famine response). In such cases they are less reliant on persuading citizens that they are not responsible.

A further strategy available to political leaders is to simply not publish any data that may reveal incompetence. However, the complete withholding of data about a politically sensitive topic can be detected by citizens, whereas the misreporting of data is harder for them to detect. In this sense the withholding of data is akin to the overt censorship of content. Both are observable. As with overt censorship, the absence of data on a salient topic may rouse the suspicions of citizens, thereby encouraging them to invest more effort into finding out about the topic (e.g. circumventing the government’s control over the internet by using virtual private networks to access blocked content) (Roberts, 2020, Chapter 4). Moreover, those efforts may lead them to uncover information about other politically sensitive topics (Hobbs & Roberts, 2018). Because the withholding of data may backfire, the fabrication of published data typically represents a less risky way for autocratic leaders to forestall collective action.

So far I have assumed that the incentive to manipulate increases if the release of accurate data will reveal bad news, with the ensuing risk of domestic criticism and protest. However, it should be noted that the incentive to falsify data also rises when a government is prescribed macroeconomic and development targets by international and regional organizations such as the International Monetary Fund, World Bank, and European Union (Aragão & Linsi, 2022; Herrera & Kapur, 2017, pp. 378–379; Sandefur & Glassman, 2015). Similarly, governments have an incentive to adjust published statistics in order to secure more funding (Kerner et al., 2017; Morgenstern, 1965, p. 21), or to demonstrate to aid donors that their money is being well spent (Jerven, 2013, pp. 75–77, 87). Autocracies may be more susceptible than democracies to both types of incentive, but in this study I will be focusing on the first. That is, controlling the flow of information so as to prevent collective action.

### How do governments manipulate reported data?

Manipulation may involve the blatant adjustment of data that has been accurately collected. This is perhaps best illustrated by the reporting of official government statistics during the Soviet era (von der Lippe 1999; Wheatcroft and Davies 1993). However, there are a range of cases where it is less obvious that the published data involves manipulation in the strict sense. Governments may, for instance, choose those estimates from a range of methodologically tenable estimates that favor its interests (Aragão and Linsi 2022; Kerner et al. 2017). The Rwandan government, for example, has been accused of deliberately underestimating income poverty during the period leading up to the 2015 referendum that made it possible for the incumbent president, Paul Kagame, to extend his rule for up to two more decades (The Economist 2019; Wilson and Blood 2019). The World Bank’s senior advisor for the region defended the estimates arguing that there is no single best way to estimate income poverty (O’Brien 2019). Similarly, some governments may have exploited the disagreement among global medical authorities during the pandemic over when to classify the death of someone infected with SARS-CoV-2 as a death due to COVID-19 (Wang et al. 2022, p. 1515). Government’s that are keen to downplay the impact of the pandemic may have erred towards reporting another cause of death in such cases (Kobak 2021). Whether these scenarios amount to manipulation may hinge on whether the reported numbers would have been adopted by a statistical agency that is free from political pressure (Aragão and Linsi 2022; Prewitt 2010). Moreover, political pressure may take the form of direct interference by the government, or self-censorship by statisticians worried about their career prospects (Jerven 2013, p. 105). Irrespective, such cases involve the systematic biasing of reported data in favor of estimates that cast the government in a favorable light in the eyes of its citizens, donors, and lenders.

It should be noted that misreporting may also take place at the sub-national level. Local officials also have an incentive to hide policy failure and demonstrate policy success to the national leaders. In China, for example, there is an incentive for local governors to overstate regional economic figures in order to advance their career prospects (Wallace 2016). The Chinese government is aware of this and adjusts its national GDP estimates downward. Nevertheless, there is some evidence that the national government has been under-correcting since 2008 (See also Angrist et al. 2021, p. 235; Chen et al. 2019).

For the purposes of this study I set aside the question of whether unelected local officials are more likely to manipulate data than elected local officials. However, it is reasonable to assume that local officials who are elected to office in an open democracy face the same disincentives as their national counterparts. That is, there is a greater risk that the misreporting of politically sensitive data will be exposed by opposition politicians, media, and civil society groups.^4^ That is not to say that democratically elected leaders do not attempt to manipulate official statistics - indeed there is evidence that it does occur, especially during periods leading up to elections (Alt et al. 2014; Gandrud and Hallerberg 2017). Rather the claim of this study is that they have more incentives to avoid fabricating official statistics and, as a result, it is less likely to occur.

### Unintentional under-reporting

The misreporting of data may occur even when a government has no intention to mislead. Thus, any analysis of data manipulation must take into account those factors independent of the incentives of political leaders that may lead to mismeasurement. At least three such factors may explain the under-reporting of mortality during the COVID-19 pandemic.

Firstly, under-reporting may be due to the capacity of the health system to tackle a novel and rapidly spreading virus. Those countries that are insufficiently prepared for such epidemics may have struggled to keep track of the number of deaths due to COVID-19, especially if their testing capacity is limited and deaths due to the pathogen are only recorded when patients have been hospitalized (Whittaker et al., 2021).

Secondly, under-reporting may arise because of a shortfall in the government’s overall capacity to collect and process policy-relevant data. Governments that lack the infrastructure necessary to collect information from all geographic locations will struggle to gather complete and accurate mortality data when faced with a rapidly spreading contagion. Information-gathering capacity is also relevant to health system capacity because the collection of infection, testing, vaccination, and mortality data is crucial for determining the right policy response at each stage of the pandemic.

Thirdly, some countries may be more vulnerable to pandemic deaths due to their epidemiological and geographic features, such as the prevalence of co-morbidities (e.g. hypertension, cardiovascular diseases, diabetes, etc.), population density, altitude, and environmental seasonality (Bollyky et al., 2022). As a result, those countries may have been overwhelmed by the rapid increase in mortality caused by the pandemic, and therefore struggled to maintain accurate mortality figures.

For example, during the first phase of the pandemic, when tests were not widely available, there was a rapid rise in deaths among the very elderly, especially in long-term care facilities. As a result, COVID-19 deaths were more likely to go unrecorded in countries with older populations (Wang et al., 2022, p. 1514).

All three of these explanations show how under-reporting can occur even when a government is not deliberately under-counting the number of deaths. Thus, it is important to control for them in order to isolate the extent to which, if at all, under-reporting is due to data manipulation.

### Estimation methods

I use ordinary least squares (OLS) regressions to assess the relationship between regime type and the manipulation of COVID-19 mortality data. The estimation model takes the following form.

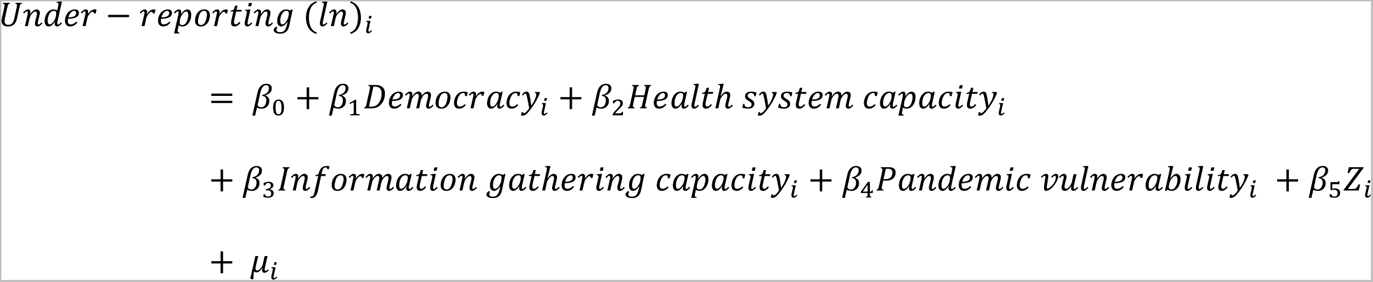

Where *Democracy* is the main independent variable of interest, *Health system capacity* is a set of variables capturing the ability of each country to respond to the pandemic, *Information-gathering capacity* is a variable capturing the ability of the government to collect and process policy-relevant data, *Pandemic vulnerability* is a set of variables capturing the extent to which each population is susceptible to COVID-19 mortality, *Z* is the set of additional control variables, and *i* is the set of countries.

I use three different indicators to estimate *under-reporting* in each country: the undercount ratio and two indicators that gauge the extent to which daily reported cases and deaths depart from expected statistical patterns (Benford-noncompliance and underdispersion). The undercount ratio is cumulative excess deaths divided by cumulative reported deaths (logged) as of December 31, 2021. The data source is the COVID-19 projections produced by the Institute for Health Metrics and Estimations (IHME) (Wang et al., 2022). IHME estimated excess deaths based on three steps. Firstly, in those locations with sufficient total mortality data, excess deaths were estimated based on a weekly or monthly comparison between mortality due to all causes and what would have been expected based on past trends and seasonality. Secondly, a statistical model was then constructed using covariates to predict the excess deaths in those locations. Thirdly, the predictive model was then used to estimate excess deaths in those locations without sufficient mortality data. In the robustness section I also test whether the baseline results hold when I use three alternative estimates of excess mortality produced by the World Health Organization (WHO, 2022a), The Economist (2020/2022), and Karlinksy and Kobak (2021).

One potential limitation of using excess mortality is that it may capture deaths that are not due to COVID-19. WHO guidelines stipulate that COVID-19 should be listed as the cause of death if a person dies with a probable or confirmed case of COVID-19, unless there is “… a clear alternative cause of death that cannot be related to COVID disease (e.g. trauma)..” (WHO, 2020). Based on this definition, the proportion of non-covid deaths captured by excess mortality is likely to be small, and that assumption is supported by two country case-studies (Wang et al., 2022, p. 1533). A related concern is that excess mortality might capture deaths that are due to contemporaneous shocks such as a natural disaster or conflict (Wang et al., 2022, p. 1534). I take steps to address this issue in the robustness section. A further consideration is that excess mortality can be negative if a country’s containment response to the pandemic also reduced deaths from other causes such as injuries and seasonal flu. Indeed five countries registered negative cumulative excess deaths by the end of 2021. I also take steps to address that issue in the robustness section below.

One advantage of measuring manipulation in terms of statistical anomalies in the reported daily COVID-19 data is that it avoids the issue of non-covid deaths and negative total deaths. Benford-noncompliance is measured in terms of the Kolmogorov-Smirnov test statistic (logged), which allows us to estimate the extent to which daily reported cases and deaths (for the period 22 January 2020 to 31 December 2021) deviate from Benford’s Law. According to that law, first digits in non-manipulated data should accord with a distribution where the number 1 is the most likely to occur and the remaining digits are increasingly less likely to occur (Tam Cho & Gaines, 2007). I provide a complete description of the steps used to measure Benford-noncompliance in the online appendix. Underdispersion is measured using the index constructed by Dimitry Kobak (2022) (logged). That index gauges the extent to which the reported cases and deaths (for the period 3 March 2020 and 30 January 2022) deviate from the expected variation in the reported cases and deaths across time. Reported COVID-19 data should fluctuate randomly across days of the week due to the nature of the data generating process. Thus, daily counts that vary smoothly across time suggest the presence of tampering.

The indicator for *democracy* is the Electoral Democracy Index from the Varieties of Democracy (V-Dem) project (Coppedge & et al, 2023). That index is scaled to a continuous interval ranging from 0 (lowest level of democracy) to 1 (highest level of democracy). It combines five components of democratic rule which aim to ensure that political leaders are sufficiently responsive to citizens: suffrage, elected officials, free and fair elections, freedom of civil and political association, and freedom of expression and access to alternative information. Those components also implicitly capture the extent to which the government’s data reporting is subject to scrutiny by media, opposition political parties, and civil society groups. If those entities are sufficiently independent of the government’s control and influence, they can collect and publish their own data and publicly question the veracity of the government’s data (World Bank, 2021, pp. 32–33, 69–70). Arguably, V-Dem’s index is methodologically superior to the other democracy indices that are currently available (Boese, 2019; Coppedge et al., 2017). Nevertheless, in the robustness section I test whether the baseline results hold when four alternative indicators of democracy are used.

*Health system capacity*, *information-gathering capacity*, and *pandemic vulnerability* capture under-reporting that is not due to deliberate under-reporting. I use two variables to capture health system capacity. GDP per capita (logged) (base 2010 international dollars) and health service capacity and access (WHO, 2021). The latter index measures the density of hospital beds and health care professionals, as well as the level of preparedness for public health events of international concern (WHO, 2019). I constructed an indicator of information-gathering capacity based on the latent factor analysis of three input variables: Hanson and Sigman’s (2021) measure of census frequency, the World Bank’s (2022) Statistical Capacity indicator, and Brambor et al’s (2020) information capacity index. I describe those input variables and the method for identifying the underlying factor in more detail in the online appendix. I use three variables to capture greater pandemic vulnerability due to factors such as seasonality, population density, and the presence of co-morbidities. These are, prevalence of lower respiratory diseases, prevalence of non-communicable diseases, and median age (GBD Collaborative Network, 2020b; UNDESA, 2022).

A further advantage of using the Benford non-compliance and underdispersion measures is that they are less likely to be affected by unintentional mismeasurement than the undercount ratio. Nevertheless, it remains possible that observed anomalies in the data are not due to an attempt by a government to mislead. For example, a government that has reached the limits of its testing capacity may in good faith estimate the numbers in a way that conflicts with the expected patterns. Thus, I retain the full set of covariates for all versions of the dependent variable.

To address the possibility of region-specific factors, I also include World Bank regions as additional control variables in all the models. All the continuous independent variables are for the year 2019 (except for median age and information-gathering capacity which are for 2015) given the likelihood that the pandemic has impacted political institutions, economic growth, the delivery of routine health care, and the spread of other respiratory pathogens. I report robust standard errors for all model specifications. Variable descriptions, summary statistics, and correlation matrices are reported in the online appendix.

### Estimations results

The results of this analysis are presented in Table 1. As we can see the democracy indicator is negatively associated with all three indicators of under-reporting – undercounting (columns 1-2), Benford non-compliance as measure by the Kolmogorov-Smirnov statistic (columns 3-5), and underdispersion (columns 6-7). Moreover, democracy remains statistically significant both with and without the covariates.

**Table 1:** Democracy and under-reporting of COVID-19.

|  | (1) | (2) | (3) | (4) | (5) | (6) | (7) |
| --- | --- | --- | --- | --- | --- | --- | --- |
| Dependent variable | Undercount ratio (IHME) (ln) | Undercount ratio (IHME) (ln) | Kolmogorov-Smirnov statistic (ln) | Kolmogorov-Smirnov statistic (ln) | Kolmogorov-Smirnov statistic (ln) | Under-dispersion index (ln) | Under-dispersion index (ln) |
| Electoral Democracy Index | -3.383***<br>(0.434) | -1.282***<br>(0.384) | -0.638***<br>(0.160) | -0.668***<br>(0.241) | -0.647**<br>(0.277) | -1.067***<br>(0.249) | -1.012***<br>(0.327) |
| GDP per capita (ln) |  | -0.387***<br>(0.127) |  | 0.174**<br>(0.0717) | 0.184**<br>(0.0783) |  | -0.211**<br>(0.0857) |
| Health service capacity & access |  | -0.0103*<br>(0.00616) |  | 0.00186<br>(0.00377) | 0.00226<br>(0.00441) |  | 0.0111**<br>(0.00521) |
| Information-gathering capacity |  | -0.256<br>(0.783) |  | 0.0495<br>(0.548) | -0.391<br>(0.615) |  | 0.422<br>(0.780) |
| Prevalence of non-communicable diseases |  | -25.57***<br>(8.552) |  | -3.019<br>(4.728) | -5.139<br>(5.518) |  | 0.472<br>(6.007) |
| Prevalence of lower respiratory diseases |  | 39.43<br>(193.5) |  | -56.53<br>(149.6) | -30.33<br>(157.2) |  | -262.7<br>(204.5) |
| Median age |  | 0.00682<br>(0.0295) |  | -0.0161<br>(0.0158) | -0.00695<br>(0.0177) |  | -0.0178<br>(0.0183) |
| Latin America & Caribbean |  | -0.982***<br>(0.281) |  | 0.619<br>(0.460) | 0.662<br>(0.470) |  | 1.516*<br>(0.839) |
| South Asia |  | -1.290***<br>(0.472) |  | 0.825<br>(0.534) | 0.864<br>(0.567) |  | 1.504*<br>(0.837) |
| Sub-Saharan Africa |  | -0.961**<br>(0.444) |  | 0.544<br>(0.493) | 0.629<br>(0.506) |  | 1.429*<br>(0.841) |
| Europe & Central Asia |  | -0.258<br>(0.205) |  | 0.537<br>(0.445) | 0.559<br>(0.455) |  | 1.794**<br>(0.821) |
| East Asia & Pacific |  | -1.701***<br>(0.514) |  | 0.424<br>(0.470) | 0.510<br>(0.488) |  | 1.572*<br>(0.821) |
| Middle East & North Africa |  | -0.625**<br>(0.292) |  | 0.399<br>(0.489) | 0.506<br>(0.510) |  | 1.867**<br>(0.836) |
| Magnitude<1000 |  |  |  | 0.461***<br>(0.122) |  |  |  |
| Constant | 4.170***<br>(0.262) | 32.25***<br>(7.791) | -2.466***<br>(0.0911) | -1.363<br>(4.183) | 0.183<br>(4.775) | 0.371***<br>(0.142) | 0.374<br>(5.398) |
| Adjusted R-squared | 0.27 | 0.709 | 0.78 | 0.165 | 0.072 | 0.103 | 0.202 |
| Observations | 171 | 168 | 169 | 166 | 135 | 167 | 166 |

Note: Undercount ratio as of December 31 2021, based on excess mortality estimated by IHME. Kolmogorov-Smirnov test statistic as of December 31 2021.
Magnitude<1000 is a dummy variable identifying those countries without at least 1000 cases or deaths on any single day. Column 5 excludes those countries from the sample. Underdispersion index as of January 30 2022. Reference category for region is North America. Robust standard errors are reported in parenthesis.
\*p<0.1; \*\*p<0.05; \*\*\*p<0.01

For the complete model using undercounting (column 2) a 10% increase in the level of democracy (i.e. a 0.1 increase in the Electoral Democracy Index’s 0-1 scale, roughly the difference between Belarus and Serbia in 2019) is associated with a 9.45% (95% CI 3.99, 14.61) reduction in the ratio. Put differently, excess mortality is predicted to be nearly ten times greater than reported mortality (9.83; 95% CI 7.35, 12.31) in countries with a democracy score of 0.25 in 2019 (e.g. Kazakhstan). By contrast, excess mortality is predicted to be nearly six times greater than reported mortality (5.98; 95% CI 5.03, 6.93) in countries with a democracy score of 0.75 in that year (e.g. Romania) (Figure 1a). For the complete model using Benford-noncompliance (column 4) a 10% increase in the level of democracy is associated with a 6.46% (95% CI 1.91, 10.81) reduction in the Kolmogorov-Smirnov statistic. Put differently, the predicted statistic for countries with democracy scores of 0.25 and 0.75 in 2019 is 0.081 (95% CI 0.07, 0.093) and 0.058 (95% CI 0.051, 0.066), respectively (Figure 1b). For the complete model using underdispersion (column 7) a 10% increase in the level of democracy is associated with a 10.59% (95% CI 4.52, 16.27) reduction in the index. Put differently, the predicted index for countries with democracy scores of 0.25 and 0.75 in 2019 is 1.41 (95% CI 1.05, 1.47) and 0.84 (95% CI 0.74, 0.94), respectively (Figure 1c).

**Figure 1:**
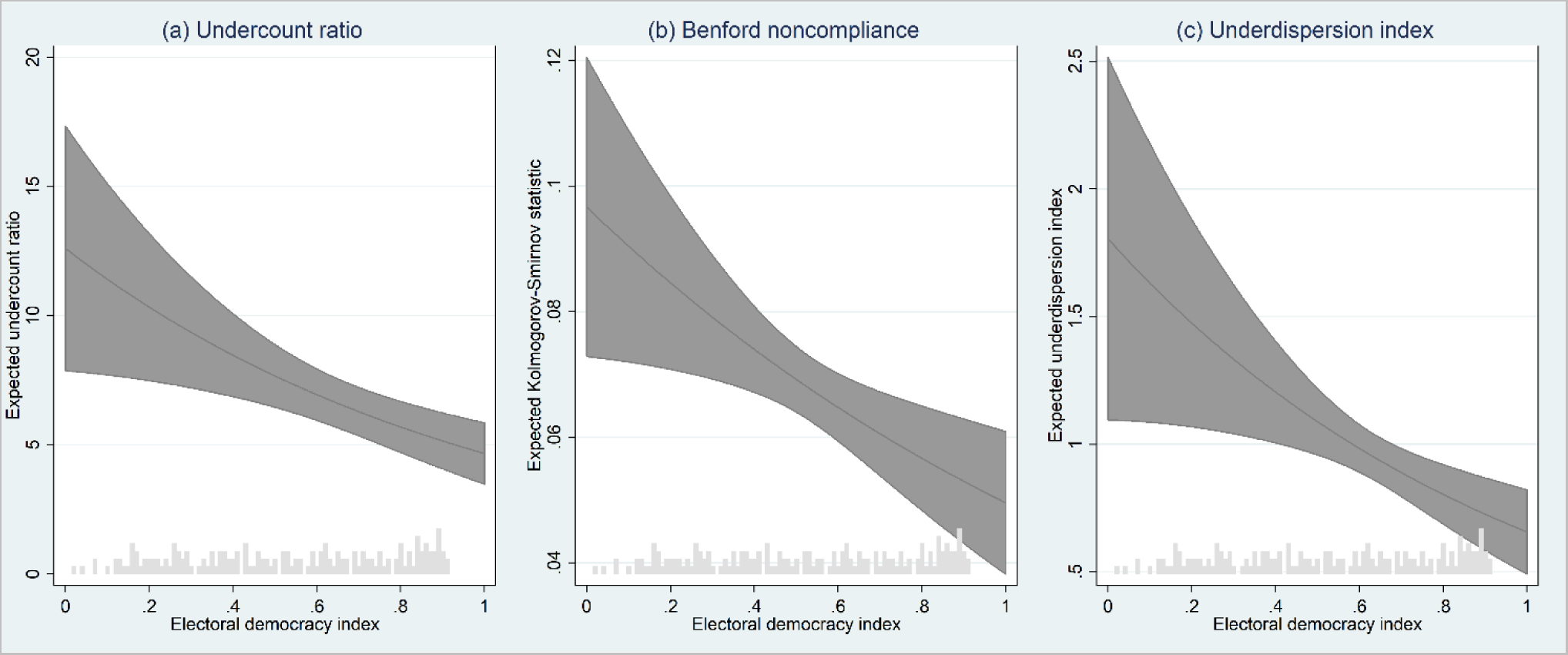
Expected under-reporting by level of democracy. *Note*: Predictions calculated with covariates set at their means. Expected values have been converted from log scale into their original scale. Shaded bands represent 95% confidence intervals. The histograms at the bottom of each figure show the frequency distribution for the democracy indicator.

Figure 2 presents the relative importance of the components that are used to construct the Electoral Democracy Index. A random forest regression was used to determine the relative predictive strength of each component for the undercount ratio. Conditional importance was used to address potential bias when strongly correlated predictors are included in the model (Strobl et al., 2008). Looking at the results in terms of the sub-components of democracy (y-axis variables), we can see that independence of opposition parties from the ruling regime is the most important predictor. Looking at the results in terms of the main components of democracy (legend), we can see that free and fair elections is the most important predictor. This suggests that data manipulation during the pandemic was more prominent in those countries where the government was able to minimize the risk of the opposition winning an election.

**Figure 2:**
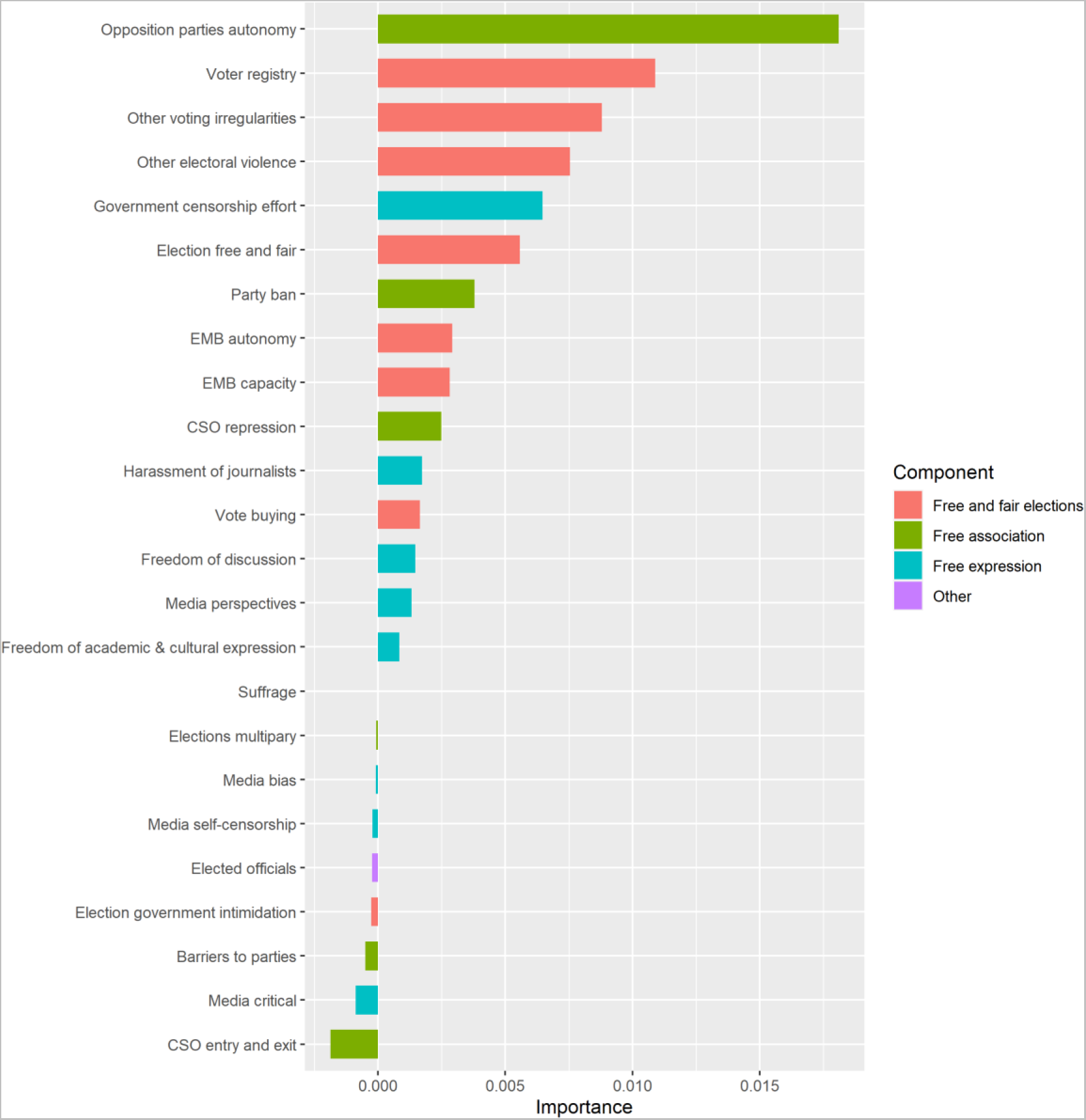
Relative importance of democracy components for undercount ratio. *Note*: The horizontal bars represent the importance (increase in mean squared error) of the 24 sub-components that are used by V-Dem to construct the five components (see legend) of the Electoral Democracy Index. All covariates included in the random forest regression model, but excluded from the figure. The R-squared for the overall model is 77.13%. All component variables are for the year 2019. EMB = electoral monitoring board. CSO = civil society organization.

### Robustness checks

I report five types of robustness check in Table 2. Firstly, I examine whether the results are affected when a dummy variable for island states is added to the set of covariates. It may be argued that island democracies such as Taiwan, Iceland, and New Zealand were blessed with a natural advantage when it came to slowing the spread of the virus. As a result, they were better placed to keep an accurate count of the number of COVID-19 deaths (column 2). Moreover, five of those island states (Australia, Iceland, New Zealand, Singapore, and Taiwan) registered negative excess deaths during the first two years of the pandemic, in part because their public health response reduced the spread of other respiratory pathogens. It is not clear whether those five countries are in some way biasing the results. Nevertheless, the addition of the dummy variable for island states provides one way to control for that possibility. I also test whether the baseline result holds when those five countries are dropped from the sample (column 3).

**Table 2:**
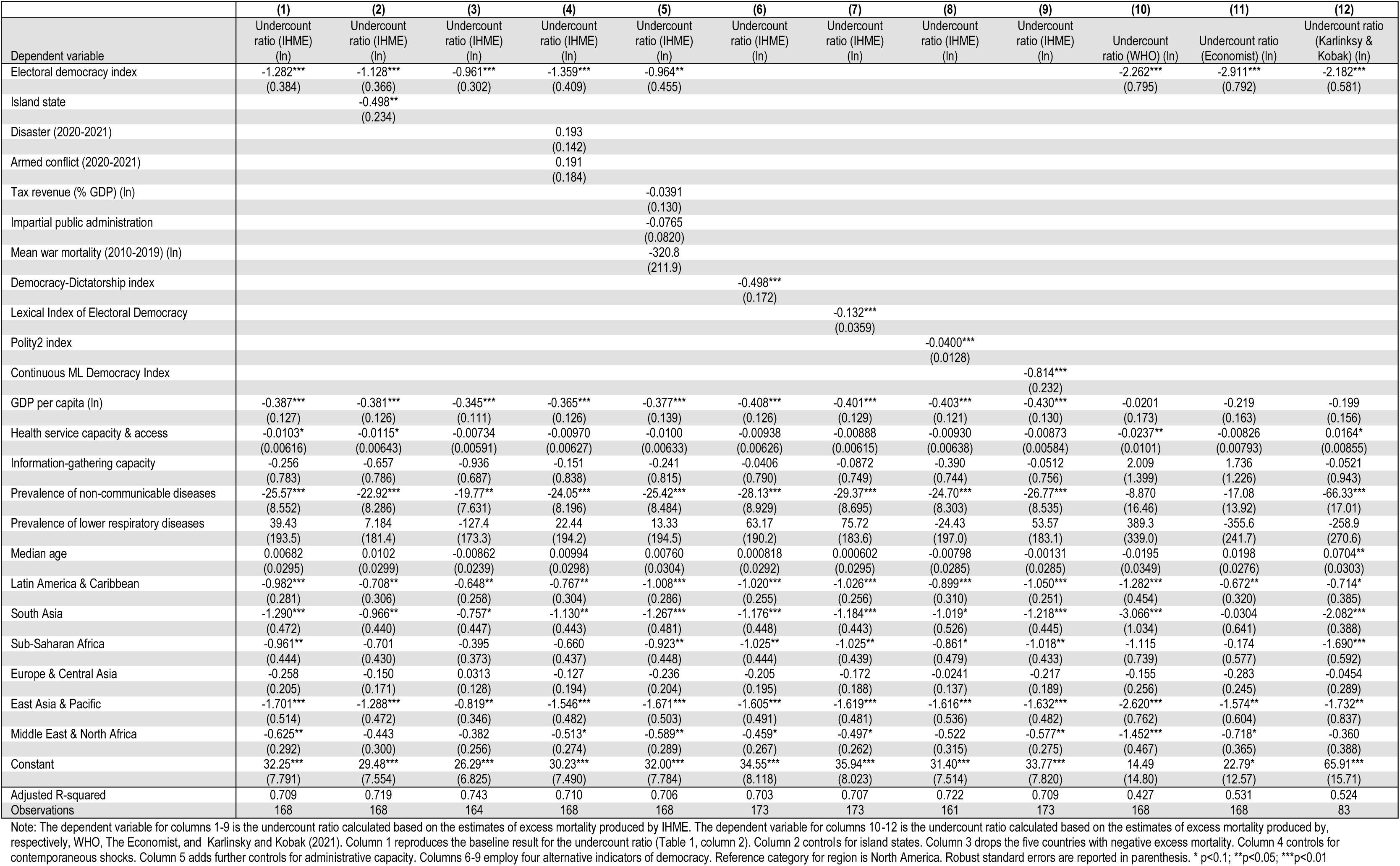
Robustness checks.

Secondly, I include dummy variables for contemporaneous disasters and conflicts (column 4). Other shocks that occurred during the pandemic may have increased excess deaths even though they are not due to COVID-19 and, therefore, do not reflect undercounting. In order to identify disasters that took place during the years 2020 and 2021 I used the Emergency Events Database compiled by the Center for Research on the Epidemiology of Disasters (Guha-Sapir, 2022). In order to identify armed conflicts that took place during those two years I used the Battle-Related Deaths Dataset (version 22.1) constructed by the Uppsala Conflict Data Program (Pettersson et al., 2021).

Thirdly, I examine whether the results hold when three indicators for administrative capacity are included among the covariates (column 5). Those indicators are tax revenue as a percentage of GDP (Heritage Foundation, n.d.) (logged), rigorous and impartial public administration (Coppedge & et al, 2023), and mean war mortality during the 10 years prior to the pandemic (GBD Collaborative Network, 2020a) (logged). It may be argued that the observed negative association between democracy and under-reporting is due to the fact that democracies are typically characterized by a greater capacity to implement policy. That is, administrative capacity, rather than democracy itself, may explain the association with under-reporting (Halleröd et al., 2013; Stasavage, 2020). Having said that, some scholars contend that the capacity to gather and process information represents a suitable proxy for administrative capacity because it is a pre-condition for the collection of taxes, as well as the successful implementation of law and policy (Brambor et al., 2020; D’Arcy & Nistotskaya, 2017; Lee & Zhang, 2017). If that is correct, then the baseline model already controls for administrative capacity. Nevertheless, I include these three further controls in order to address the possibility that information-gathering capacity, by itself, does not fully capture overall administrative capacity.

Fourthly, I examine whether the results hold when four alternative indicators of democracy are used (columns 6-9). Specifically, the dichotomous Democracy-Dictatorship index (Bjørnskov & Rode, 2020), the Lexical Index of Electoral Democracy (Skaaning et al., 2015), the polychotomous Polity2 index (Marshall & Gurr, 2020), and the continuous Machine Learning (ML) Democracy Index (Gründler & Krieger, 2021). These indicators, along with V-Dem’s Electoral Democracy Index, represent distinct ways to conceptualize and measure the level of democracy in each country. Thus, it is important to check whether the baseline results are sensitive to the selection of democracy indicator.

Fifthly, I examine whether the results hold when undercounting is calculated based on the estimates of excess deaths produced by World Health Organization (WHO, 2022a), The Economist (2020/2022), and Karlinksy and Kobak (2021) (columns 10-12). It is important to check whether the baseline results are not merely an artifact of the particular estimation method used by IHME. As before, I use cumulative excess deaths up to the end of December 2021. Like IHME, WHO and The Economist used covariate prediction models based on those countries with sufficient all-cause mortality data to generate estimates for those locations without sufficient data.^5^ However, it may be argued that the prediction models used by those three organizations are built based on countries whose characteristics do not adequately represent the countries for which they aim to provide estimates (Adam, 2022). One advantage of Karlinksy and Kobak’s estimates is that they are restricted to the 101 countries with sufficiently complete vital statistics, and so they are not model dependent. This comes at cost, however, because the excluded countries may be self-selecting based on regime type.

For ease of comparison the first column in Table 2 replicates the baseline results for the undercount ratio (Table 1, column 2). As we can see each of the robustness checks is consistent with the baseline finding. Nevertheless, I cannot rule out the possibility that there are other factors that may explain the association between regime type and the under-reporting of COVID-19.

## Discussion

These results indicate that democracy remains negatively associated with under-reporting after controlling for pre-existing characteristics that may affect each government’s ability to collect accurate COVID-19 data. This in turn implies that autocratic leaders are more likely to manipulate COVID-19 data than their democratic counterparts. However, even with the inclusion of a number control variables and a range of robustness checks, it remains possible that omitted factors are driving the results. Thus, these results should be seen as providing suggestive, rather than conclusive, evidence.

Nevertheless, they are consistent with existing research on the way in which the political survival of autocrats is dependent on their ability to manage the information available to citizens (Carlitz & McLellan, 2021; Hollyer et al., 2015; King et al., 2013; Little, 2017; Lorentzen, 2014; Stockmann & Gallagher, 2011). Indeed there is growing evidence that the balance between information control and repression in autocracies has shifted over the last two decades. The new breed of autocrat places more emphasis on the manipulation of information than the inculcation of fear in order to prolong their tenure in power (Guriev & Treisman, 2019). According to that approach to regime survival, the key is to prevent disgruntled citizens from becoming aware that there are a sufficient number of them to overthrow the government. The threat of repression remains as a deterrent, but that may not suffice if a critical number of citizens manage to overcome the collective action problem. Moreover, traditional repression is more likely to attract the attention of the international community, raising the prospect of sanctions and the withdrawal of financial aid. I have argued that the misreporting of policy-relevant statistics remains an important means for the autocrat to block access to the information necessary for collective action, even as they endeavor to persuade citizens that they are not responsible for bad outcomes. The autocrat’s own lack of information about the degree to which citizens believe their interpretation of the bad news, means they prefer to also hide the amount of bad news.

During the pandemic at least two authoritarian states adopted a different approach to information control. Rather than undercounting the number of deaths, the governments of Tanzania and Turkmenistan simply denied the presence of the virus in their countries (Human Rights Watch, 2021; Mwai & Giles, 2021). Denial precluded the very need to report data or to spin bad news. However, even though it is difficult for citizens to ascertain the exact death toll, it would have been increasingly obvious that a deadly contagion was spreading through their communities. Persuading citizens that the death count is lower than elsewhere would have become an easier proposition than persuading them that there are no deaths to count in the first place. Moreover, denial would have made it very difficult to implement public health policies designed to limit the number of cases and deaths. Unsurprisingly, therefore, nearly all other governments chose to regularly release mortality data during the pandemic. The Tanzanian government did eventually report some mortality data, but the number of data releases were few and far between, and likely severely understated the true death toll (by the end of 2021, for example, reported deaths in that country were 180 times lower than the excess mortality estimated by IHME). Generally speaking, the withholding of economic and development data is now less common than it used to be due to the expectations of international organizations, aid donors, and investors (Carlitz & McLellan, 2021). Moreover, because the complete withholding of information is observable it may raise suspicions among citizens, thereby encouraging them to seek out more information about the topic the government wishes to hide (Roberts, 2020, Chapter 4). However, while governments have an incentive to release policy-relevant statistics, the incentive to doctor them remains. Paying lip service to transparency while at that same time publishing falsified information represents a more nuanced and potentially more successful way for autocrats to prevent collective action.

One area that the current study does not fully explore is the extent to which deliberate under-reporting is due to the revision of data received by the national government, or conscious attempts by the government to prevent the collection of accurate data in the first place (e.g. deliberately curtailing COVID-19 testing, such that it is harder for healthcare workers to assign the cause of death in each case). Autocratic leaders are likely to prefer the first approach because accurate information is often needed in order to develop an adequate policy response and, thereby, to forestall criticism and protest. However, it remains possible that mortality numbers received by the national government are being deliberately underestimated by local officials, keen to hide their failure in handling the pandemic from their superiors (Wallace, 2016). In other words, it may be the national government itself that is being misled. However, this less likely to occur in a democratic context because local officials themselves are exposed to scrutiny from the opposition, media, and civil society. Irrespective, the regression results for statistical irregularities - Benford-noncompliance and underdispersion - suggest that autocratic governments, at a minimum, manipulate data that has already been collected at the national level.

It might be argued that citizens care more about economic outcomes than population health outcomes, or that they are more likely to hold governments’ responsible for bad economic outcomes than bad health outcomes. Indeed there is some evidence that citizens in democracies are already pre-disposed to treat pandemics as natural phenomena that are beyond the control of policy-makers (Acharya et al., 2020; Achen & Bartels, 2017, pp. 140– 142). If that is correct then there is less incentive for political leaders to manipulate population health data. The results of this study suggest that, even if that is the case, there remains an incentive for autocratic leaders to misinform citizens about their health risks and status.

Finally, it should be noted that the results presented here relate to data manipulation after an epidemiological shock. These findings likely apply to the propensity for manipulation following other kinds of shock such as war, famine, natural disaster, or severe economic recession. Arguably, the threat to the government’s survival is less pronounced in the absence of severe shocks, and so the incentive to falsify data is reduced in such cases. Still, existing research indicates that autocrats fabricate economic data even in the absence of a recession (Carlitz & McLellan, 2021; Magee & Doces, 2015; Martínez, 2022). Nevertheless, further research is needed to determine whether data relating to adverse health outcomes that are not due to a shock are more likely to be manipulated by autocrats.

## Conclusion

It is fundamentally difficult to assess the relationship between regime type and data manipulation because researchers typically do not have access to statistics from sources that are not controlled by the government. The COVID-19 pandemic represents a unique opportunity to examine that relationship because of its widespread impact and the ability to compare reported deaths with excess deaths and examine reported cases and deaths for statistical irregularities. Using estimates of excess mortality for a large number of countries, I find evidence that autocratic governments are more likely to deliberately undercount deaths due to the pathogen than their democratic counterparts. Similarly, I find that the case and death counts reported by autocratic regimes are more likely to feature statistical anomalies. These results hold when controls are included for unintentional mismeasurement and after running a battery of robustness checks. Overall, these results suggest that autocratic leaders manipulate data that may trigger criticism and protest, even though they can use their control over traditional and online media to disown responsibility for bad news.

This conclusion is consistent with two previous cross-national studies on the association between regime type and the manipulation of national income statistics (Magee & Doces, 2015; Martínez, 2022). Taken together these three studies imply that politically-sensitive data are systematically biased in favor of autocratic regimes. If this general finding is confirmed by subsequent research, it presents a significant problem for citizens, researchers, and international organizations. Firstly, the absence of accurate information may prevent citizens from being able to make the decisions necessary to protect their own well-being (e.g. governments that deliberately understate the threat posed by a disease limit the ability of individuals to take steps to avoid preventable morbidity and premature mortality). Secondly, aggregate indices of economic and human development may overstate the performance of autocratic regimes if the input data are directly sourced from each government. Thirdly, cross-national studies that examine the association between regime type and policy outcomes such as economic growth, educational attainment, and infant mortality may be biased in favor of autocratic regimes (Knutsen, 2021, p. 1509). Fourthly, the manipulation of data makes it harder for international organizations and aid donors to determine whether recommended targets have been achieved and whether they are supporting the right policies.

One solution to these problems is to rely on proxy indicators, or the covariate model approach outlined in this study, to estimate the outcome of interest in those cases where the reported data are suspect. Reported data may be deemed to be questionable in those cases where there are unexplained statistical anomalies, or the national statistical agency is subject to the direct influence of the government.^6^ The latter suggests a more forward-looking approach. Namely, the advocacy of reforms designed to ensure the institutional independence of statistical agencies (Taylor 2016, pp. 15–20). There is a growing emphasis on building the capacity of such agencies (e.g. Dang et al. 2021), but so far less weight has been placed on reforms designed to minimize their exposure to political interference. This stands in stark contrast with well-established efforts to promote the independence of central banks (Herrera and Kapur 2017, p. 375; Jolliffe et al. 2021).

## Data Availability

All data produced in the present study and replication codes are available upon reasonable request to the author

## Footnotes

1 In that context it is typically very difficult to find a suitable proxy indicator to estimate the actual level of morbidity and mortality. Two studies have attempted to estimate actual pandemic deaths by using satellite imagery of grave sites in cemeteries (Koum Besson et al. 2021; Warsame et al. 2021). However, that approach cannot be used in those countries where cremation is commonly practiced and, unlike night time lights, political leaders can take steps to hide the evidence. Thus, this approach is better reserved for assisting governments that lack the capacity to collect accurate mortality data for themselves, rather than as a means to detect deliberate undercounting.

2 Crucially an autocratic government need not solely rely on state media to propagate its interpretation of events. Instead it may disguise its involvement by recruiting individuals to repeat its interpretation on social media, or via news outlets that it does not directly control. This approach enables the government to crowd out interpretations that imply policy failure, thereby reducing the need to directly censor content. The upshot of this is that narrative control can take place even when citizens have access to multiple sources of information (Roberts 2020, Chapter 6).

3 Manipulation that is clearly inconsistent with the experienced reality of citizens is both observable and ineffective (Cavallo et al., 2016). In addition, it may reduce citizens’ trust in official statistics in general, including those that have not been manipulated (e.g. citizens may interpret accurately reported epidemic deaths as underestimates if the government has a track record of understating inflation statistics). Nevertheless, autocrats may still engage in observable data manipulation because it enables them to signal strength and, thereby, deter dissent (Huang, 2015).

4 Some scholars contend that the absence of information checks is the Achilles heel of autocratic rule (Egorov et al. 2009; Sen 1999, pp. 180–182; Wigley and Akkoyunlu-Wigley 2017). Autocratic leaders typically lack the means to independently verify the information provided by local officials and, therefore, cannot be sure whether they have selected the right policies, or whether those officials are correctly implementing policy. The consequences of this blind spot can be catastrophic. During the Great Leap Forward in China lower-level bureaucrats over-reported grain production leading the central government to extract too much for urban centers, leaving farmers with insufficient food (Wallace 2016). This contributed to the emergence of a famine that killed an estimated 30 million people in China between 1959 and 1961.

5 Nevertheless, there is some important differences between the three estimation projects in terms of the precise model design, covariates selection, and data sources for all-cause mortality. All three projects provide detailed descriptions of their methodologies (The Economist, 2021; Wang et al., 2022; WHO, 2022b). In addition, IHME and The Economist have made their replication code publicly available (The Economist, 2020/2022; Wang & & et al, 2022/2022).

6 Two existing datasets provide indicators of the extent to which national statistical agencies are free from political influence. Angrist et al (2021) have extracted three binary indicators of independence from IMF audit data for 79 countries. The Ibrahim Index of African Governance (Mo Ibrahim Foundation n.d.) includes an indicator that captures the independence of statistical agencies in that region. Ideally, a fine-grained indicator with global coverage would be developed, along the lines of the one produced for the Ibrahim Index and the one for electoral monitoring board autonomy produced by the V-Dem project (Coppedge and et al 2022).

## References

1. Acharya, A., Gerring, J., & Reeves, A. (2020). Is health politically irrelevant? Experimental evidence during a global pandemic. BMJ Global Health, 5(10), e004222. https://doi.org/10.1136/bmjgh-2020-004222

2. Achen, C. H., & Bartels, L. M. (2017). Democracy for Realists: Why Elections Do Not Produce Responsive Government (Revised edition). Princeton University Press.

3. Adam, D. (2022). The pandemic’s true death toll: Millions more than official counts. Nature, 601(7893), 312–315. https://doi.org/10.1038/d41586-022-00104-8

4. Aragão, R., & Linsi, L. (2022). Many shades of wrong: What governments do when they manipulate statistics. Review of International Political Economy, 29(1), 88–113. https://doi.org/10.1080/09692290.2020.1769704

5. Ashton, B., Hill, K., Piazza, A., & Zeitz, R. (1984). Famine in China, 1958-61. Population and Development Review, 10(4), 613–645. https://doi.org/10.2307/1973284

6. Bjørnskov, C., & Rode, M. (2020). Regime types and regime change: A new dataset on democracy, coups, and political institutions. The Review of International Organizations, 15(2), 531–551. https://doi.org/10.1007/s11558-019-09345-1

7. Boese, V. A. (2019). How (not) to measure democracy. International Area Studies Review, 22(2), 95–127. https://doi.org/10.1177/2233865918815571

8. Bollyky, T. J., Hulland, E. N., Barber, R. M., Collins, J. K., Kiernan, S., Moses, M., Pigott, D. M., Jr, R. C. R., Sorensen, R. J. D., Abbafati, C., Adolph, C., Allorant, A., Amlag, J. O., Aravkin, A. Y., Bang-Jensen, B., Carter, A., Castellano, R., Castro, E., Chakrabarti, S., … Dieleman, J. L. (2022). Pandemic preparedness and COVID-19: An exploratory analysis of infection and fatality rates, and contextual factors associated with preparedness in 177 countries, from Jan 1, 2020, to Sept 30, 2021. The Lancet, 0(0). https://doi.org/10.1016/S0140-6736(22)00172-6

9. Brambor, T., Goenaga, A., Lindvall, J., & Teorell, J. (2020). The Lay of the Land: Information Capacity and the Modern State. Comparative Political Studies, 53(2), 175–213. https://doi.org/10.1177/0010414019843432

10. Brewer, J. (1990). The sinews of power: War, money, and the English state, 1688-1783. Harvard University Press.

11. Carlitz, R. D., & McLellan, R. (2021). Open Data from Authoritarian Regimes: New Opportunities, New Challenges. Perspectives on Politics, 19(1), 160–170. https://doi.org/10.1017/S1537592720001346

12. Cavallo, A., Cruces, G., & Perez-Truglia, R. (2016). Learning from Potentially Biased Statistics. Brookings Papers on Economic Activity, 59–92.

13. Chen, W., Chen, X., Hsieh, C.-T., & Song, Z. (2019). A Forensic Examination of China’s National Accounts. Brookings Papers on Economic Activity, 77–141.

14. Coppedge, M., &, et al. (2023). V-Dem [Country-Year/Country-Date] Dataset v13 (Version 13) [Data set]. Varieties of Democracy Institute. https://doi.org/10.23696/vdemds23.

15. Coppedge, M., Gerring, J., Lindberg, S. I., Skaaning, S.-E., & Teorell, J. (2017). V-Dem Comparisons and Contrasts with Other Measurement Projects (Working Paper No. 45). Varieties of Democracy (V-Dem) Institute. http://hdl.handle.net/2077/52225

16. D’Arcy, M., & Nistotskaya, M. (2017). State First, Then Democracy: Using Cadastral Records to Explain Governmental Performance in Public Goods Provision. Governance, 30(2), 193–209. https://doi.org/10.1111/gove.12206

17. GBD Collaborative Network. (2020a). Global Burden of Disease Study 2019 (GBD 2019) Covariates 1980-2019 (Global Burden of Disease) [Data set]. Institute for Health Metrics and Evaluation (IHME), University of Washington. https://doi.org/10.6069/CFCY-WA51

18. GBD Collaborative Network. (2020b). Global Burden of Disease Study 2019 (GBD 2019) Results. [Data set]. Institute for Health Metrics and Evaluation (IHME), University of Washington. http://ghdx.healthdata.org/gbd-results-tool

19. Gründler, K., & Krieger, T. (2021). Using Machine Learning for measuring democracy: A practitioners guide and a new updated dataset for 186 countries from 1919 to 2019. European Journal of Political Economy, 70, 102047. https://doi.org/10.1016/j.ejpoleco.2021.102047

20. Guha-Sapir, D. (2022). EM-DAT. The Emergency Events Database [Data set]. Université Catholique de Louvain (UCL) - CRED. https://www.emdat.be/

21. Guriev, S., & Treisman, D. (2019). Informational Autocrats. Journal of Economic Perspectives, 33(4), 100–127. https://doi.org/10.1257/jep.33.4.100

22. Halleröd, B., Rothstein, B., Daoud, A., & Nandy, S. (2013). Bad Governance and Poor Children: A Comparative Analysis of Government Efficiency and Severe Child Deprivation in 68 Low- and Middle-income Countries. World Development, 48, 19–31. https://doi.org/10.1016/j.worlddev.2013.03.007

23. Heritage Foundation. (n.d.). Economic Data and Statistics on World Economy and Economic Freedom [Dataset]. Heritage Foundation. Retrieved March 26, 2021, from https://www.heritage.org/index/download

24. Herrera, Y. M., & Kapur, D. (2017). Improving Data Quality: Actors, Incentives, and Capabilities. Political Analysis, 15(4), 365–386. https://doi.org/10.1093/pan/mpm007

25. Hobbs, W. R., & Roberts, M. E. (2018). How Sudden Censorship Can Increase Access to Information. American Political Science Review, 112(3), 621–636. https://doi.org/10.1017/S0003055418000084

26. Hollyer, J. R., Rosendorff, B. P., & Vreeland, J. R. (2011). Democracy and Transparency. The Journal of Politics, 73(04), 1191–1205. https://doi.org/10.1017/S0022381611000880

27. Hollyer, J. R., Rosendorff, B. P., & Vreeland, J. R. (2015). Transparency, Protest, and Autocratic Instability. American Political Science Review, 109(4), 764–784. https://doi.org/10.1017/S0003055415000428

28. Huang, H. (2015). Propaganda as Signaling. Comparative Politics, 47(4), 419–444. https://doi.org/10.5129/001041515816103220

29. Human Rights Watch. (2021). Turkmenistan: Events of 2021. In *World Report* 2022. https://www.hrw.org/world-report/2022/country-chapters/turkmenistan

30. Jerven, M. (2013). Poor Numbers: How We Are Misled by African Development Statistics and What to Do about It. Cornell University Press.

31. Jiang, J., & Yang, D. L. (2016). Lying or Believing? Measuring Preference Falsification From a Political Purge in China. Comparative Political Studies, 49(5), 600–634. https://doi.org/10.1177/0010414015626450

32. Karlinsky, A., & Kobak, D. (2021). Tracking excess mortality across countries during the COVID-19 pandemic with the World Mortality Dataset. ELife, 10, e69336. https://doi.org/10.7554/eLife.69336

33. Kerner, A., Jerven, M., & Beatty, A. (2017). Does it pay to be poor? Testing for systematically underreported GNI estimates. The Review of International Organizations, 12(1), 1–38. https://doi.org/10.1007/s11558-015-9239-3

34. Kilani, A. (2021). Authoritarian regimes’ propensity to manipulate Covid-19 data: A statistical analysis using Benford’s Law. Commonwealth & Comparative Politics, 59(3), 319–333. https://doi.org/10.1080/14662043.2021.1916207

35. King, G., Pan, J., & Roberts, M. E. (2013). How Censorship in China Allows Government Criticism but Silences Collective Expression. American Political Science Review, 107(02), 326–343. https://doi.org/10.1017/S0003055413000014

36. Knutsen, C. H. (2021). A business case for democracy: Regime type, growth, and growth volatility. Democratization, 28(8), 1505–1524. https://doi.org/10.1080/13510347.2021.1940965

37. Knutsen, C. H., & Kolvani, P. (2022). Fighting the Disease or Manipulating the Data? Democracy, State Capacity, and the COVID-19 Pandemic (Working Paper No. 127). Varieties of Democracy Institute.

38. Kobak, D. (2022). Underdispersion: A statistical anomaly in reported Covid data. Significance, 19(2), 10–13. https://doi.org/10.1111/1740-9713.01627

39. Lamberova, N., & Sonin, K. (2022). Information Manipulation and Repression: A Theory and Evidence from the COVID Response in Russia (Working Paper No. 2022–101; Becker Friedman Institute). University of Chicago. https://bfi.uchicago.edu/working-paper/information-manipulation-and-repression-a-theory-and-evidence-from-the-covid-response-in-russia/

40. Lee, M. M., & Zhang, N. (2017). Legibility and the Informational Foundations of State Capacity. The Journal of Politics, 79(1), 118–132. https://doi.org/10.1086/688053

41. Little, A. T. (2017). Propaganda and credulity. Games and Economic Behavior, 102, 224–232. https://doi.org/10.1016/j.geb.2016.12.006

42. Lorentzen, P. L. (2014). China’s Strategic Censorship. American Journal of Political Science, 58(2), 402–414. https://doi.org/10.1111/ajps.12065

43. Magee, C. S. P., & Doces, J. A. (2015). Reconsidering Regime Type and Growth: Lies, Dictatorships, and Statistics. International Studies Quarterly, 59(2), 223–237.

44. Marshall, M. G., & Gurr, T. R. (2020). Polity5: Political Regime Characteristics and Transitions, 1800-2018 (Data Set Polity5 dataset version 2018). Center for Systemic Peace. http://www.systemicpeace.org/inscrdata.html

45. Martínez, L. R. (2022). How Much Should We Trust the Dictator’s GDP Growth Estimates? Journal of Political Economy, 000–000. https://doi.org/10.1086/720458

46. Michalski, T., & Stoltz, G. (2013). Do Countries Falsify Economic Data Strategically? Some Evidence That They Might. The Review of Economics and Statistics, 95(2), 591–616.

47. Morgenstern, O. (1965). On the Accuracy of Economic Observations (2nd Edition). Princeton University Press.

48. Mwai, P., & Giles, C. (2021, March 17). Covid: Does Tanzania have a hidden epidemic? BBC News. https://www.bbc.com/news/56242358

49. Neumayer, E., & Plümper, T. (2022). Does ‘Data fudging’ explain the autocratic advantage? Evidence from the gap between Official Covid-19 mortality and excess mortality. SSM - Population Health, 19, 101247. https://doi.org/10.1016/j.ssmph.2022.101247

50. Pettersson, T., Davies, S., Deniz, A., Engström, G., Hawach, N., Högbladh, S., & Öberg, M. S. M. (2021). Organized violence 1989–2020, with a special emphasis on Syria. Journal of Peace Research, 58(4), 809–825. https://doi.org/10.1177/00223433211026126

51. Roberts, M. E. (2020). Censored: Distraction and Diversion Inside China’s Great Firewall (Reprint edition). Princeton University Press.

52. Rozenas, A., & Stukal, D. (2019). How Autocrats Manipulate Economic News: Evidence from Russia’s State-Controlled Television. The Journal of Politics, 81(3), 982–996. https://doi.org/10.1086/703208

53. Sandefur, J., & Glassman, A. (2015). The Political Economy of Bad Data: Evidence from African Survey and Administrative Statistics. The Journal of Development Studies, 51(2), 116–132. https://doi.org/10.1080/00220388.2014.968138

54. Schedler, A. (2013). The Politics of Uncertainty: Sustaining and Subverting Electoral Authoritarianism. Oxford University Press. https://doi.org/10.1093/acprof:oso/9780199680320.001.0001

55. Skaaning, S.-E., Gerring, J., & Bartusevičius, H. (2015). A Lexical Index of Electoral Democracy. Comparative Political Studies, 48(12), 1491–1525. https://doi.org/10.1177/0010414015581050

56. Spagat, M., & van Weezel, S. (2017). Half a million excess deaths in the Iraq war: Terms and conditions may apply. Research & Politics, 4(4), 2053168017732642. https://doi.org/10.1177/2053168017732642

57. Stasavage, D. (2020). Democracy, Autocracy, and Emergency Threats: Lessons for COVID-19 From the Last Thousand Years. International Organization, 74(S1), E1–E17. https://doi.org/10.1017/S0020818320000338

58. Stockmann, D., & Gallagher, M. E. (2011). Remote Control: How the Media Sustain Authoritarian Rule in China. Comparative Political Studies, 44(4), 436–467. https://doi.org/10.1177/0010414010394773

59. Strobl, C., Boulesteix, A.-L., Kneib, T., Augustin, T., & Zeileis, A. (2008). Conditional variable importance for random forests. BMC Bioinformatics, 9(1), 307. https://doi.org/10.1186/1471-2105-9-307

60. Tam Cho, W. K., & Gaines, B. J. (2007). Breaking the (Benford) Law. The American Statistician, 61(3), 218–223. https://doi.org/10.1198/000313007X223496

61. The Economist. (2022). The Economist’s tracker for covid-19 excess deaths [Data set & code]. The Economist. https://github.com/TheEconomist/covid-19-excess-deaths-tracker (Original work published 2020)

62. The Economist. (2021, May 13). How we estimated the true death toll of the pandemic. *The Economist*. https://www.economist.com/graphic-detail/2021/05/13/how-we-estimated-the-true-death-toll-of-the-pandemic

63. UNDESA. (2022). World Population Prospects 2022 [Data set]. United Nations Department of Economic and Social Affairs. https://population.un.org/wpp/

64. Viboud, C., Simonsen, L., Fuentes, R., Flores, J., Miller, M. A., & Chowell, G. (2016). Global Mortality Impact of the 1957–1959 Influenza Pandemic. The Journal of Infectious Diseases, 213(5), 738–745. https://doi.org/10.1093/infdis/jiv534

65. Wallace, J. L. (2016). Juking the Stats? Authoritarian Information Problems in China. British Journal of Political Science, 46(1), 11–29. https://doi.org/10.1017/S0007123414000106

66. Wallace, J. L. (2022). Seeking Truth and Hiding Facts: Information, Ideology, and Authoritarianism in China. Oxford University Press.

67. Wang, H., & &, et al. (2022). Code for estimating excess mortality due to the COVID-19 pandemic: A systematic analysis of COVID-19-related mortality, 2020–21 [R]. IHME Demographics. https://github.com/ihmeuw-demographics/publication_covid_em (Original work published 2022)

68. Wang, H., Paulson, K. R., Pease, S. A., Watson, S., Comfort, H., Zheng, P., Aravkin, A. Y., Bisignano, C., Barber, R. M., Alam, T., Fuller, J. E., May, E. A., Jones, D. P., Frisch, M. E., Abbafati, C., Adolph, C., Allorant, A., Amlag, J. O., Bang-Jensen, B., … Murray, C. J. L. (2022). Estimating excess mortality due to the COVID-19 pandemic: A systematic analysis of COVID-19-related mortality, 2020–21. The Lancet, 399(10334), 1513–1536. https://doi.org/10.1016/S0140-6736(21)02796-3

69. Wedeen, L. (2015). Ambiguities of Domination: Politics, Rhetoric, and Symbols in Contemporary Syria. University of Chicago Press.

70. Whittaker, C., Walker, P. G. T., Alhaffar, M., Hamlet, A., Djaafara, B. A., Ghani, A., Ferguson, N., Dahab, M., Checchi, F., & Watson, O. J. (2021). Under-reporting of deaths limits our understanding of true burden of covid-19. BMJ, 375, n2239. https://doi.org/10.1136/bmj.n2239

71. WHO. (2019). Primary health care on the road to universal health coverage: 2019 global monitoring report (9789240029040 (electronic version); p. 5). World Health Organization. https://apps.who.int/iris/handle/10665/328913

72. WHO. (2020). International Guidelines for Certification and Classification (Coding) of COVID-19 as Cause of Death. World Health Organization. https://www.who.int/publications/m/item/international-guidelines-for-certification-and-classification-(coding)-of-covid-19-as-cause-of-death

73. WHO. (2021). Index of service capacity and access (Global Health Observatory) [Data set]. World Health Organization. https://www.who.int/data/gho/data/indicators/indicator-details/GHO/uhc-sci-components-service-capacity-and-access

74. WHO. (2022a). Global excess deaths associated with COVID-19 (modelled estimates) [Data set]. World Health Organization. https://www.who.int/data/sets/global-excess-deaths-associated-with-covid-19-modelled-estimates

75. WHO. (2022b). Methods for estimating the excess mortality associated with the COVID-19 pandemic. World Health Organization. https://www.who.int/publications/m/item/methods-for-estimating-the-excess-mortality-associatedwith-the-covid-19-pandemic

76. World Bank (Ed.). (2021). World Development Report 2021: Data for better lives. World Bank.

77. World Bank. (2022). Statistical Capacity Indicator [Data set]. World Bank. https://datatopics.worldbank.org/statisticalcapacity/

