## Appendix for "Regime type and Data Manipulation: Evidence from the COVID-19 Pandemic"

### Online appendix for ‘Regime-type and Data Manipulation: Evidence from the COVID-19 Pandemic’

Simon Wigley

Bilkent University

### 1. Variable descriptions and sources

| Variable | Description | Period | Source(s) |
|---|---|---|---|
| **Undercount ratio (ln) (IHME)** | Ratio of cumulative excess deaths due to COVID-19 (IHME estimates) over cumulative reported deaths due to COVID-19 | January 2020- December 31 2021 | Wang et al (2022) and Dong et al (2020) |
| **Benford-noncompliance (ln)** | Measures the extent to which first digit frequencies deviate from the frequencies that would be expected based on Benford's Law. Measured in terms of the Kolmogorov-Smirnov test statistic. | 23 January 2020 - 31 December 2021 | Calculated by author based on reported daily cases and deaths from Dong et al (2020). (See complete description in section 7 below) |
| **Underdispersion index (ln)** | Measures the extent to which the reported cases and deaths (for the period) deviate from the expected variation in the reported cases and deaths across time. | 3 March 2020- 30 January 2022 | Kobak (2022) (variable name: Underdispersion ratio (max)) |
| **Electoral Democracy Index** | Captures level of democracy. Combines scores for suffrage, free and fair elections, elected officials, freedom of civil and political organization, and freedom of expression. | 2019 | Varieties of Democracy (V-Dem) Project (Coppedge and et al 2023) (variable name: v2x_polyarchy) |
| **GDP per capita (ln)** | GDP per capita in base 2010 international dollars. | 2019 | GBD Collaborative Network (2020a) (variable name: GDPpc_id_b2010) |
| **Health service capacity & access index** | Captures the density of hospital beds and health care professionals, as well as the level of preparedness for public health events of international concern | 2019 | WHO (2021) (Variable name: UHC_SCI_CAPACITY) |
| **Information-gathering capacity** | Captures the ability of the government to collect and process data from all regions within its territory. | 2015 | Constructed by the author using input variables from Hanson and Sigman (2021), the World Bank (2022), and Brambor et al (2020). (see complete description in section 8 below) |
| **Prevalence of lower respiratory diseases** | Percentage of the population with upper or lower respiratory diseases (all ages) | 2019 | GBD Collaborative Network (2020b) |
| **Prevalence of non-communicable diseases** | Percentage of the population with non-communicable diseases (all ages) | 2019 | GBD Collaborative Network (2020b) |
| **Median age** | Population median age (years) | 2015 | UNDESA (2022) |
| **Region** | World Bank regions: Europe and Central Asia, Sub-Saharan Africa, Middle East and North Africa, South Asia, East Asia and Pacific, Latin America and Caribbean, North America | Fixed factor | World Bank (n.d.) |
| **Island states** | States without a land border. | Fixed factor | Author |
| **Disaster** | Natural or technological disaster events during 2020 or 2021 that caused deaths per capita beyond the 50$^{th}$ percentile for all such events during those years, or affected more people per capita than the 90$^{th}$ percentile for all such events during those years. The latter was included because (1) less lethal disasters that affected a large proportion of the population would have compromised the ability of the government to respond to the pandemic and collect accurate data, and (2) some of these events are recent and so death counts may not be currently available. | 2020-2021 | Constructed by the author based on The Emergency Events Database (EM-DAT) (Guha-Sapir 2022) |

| | | | |
|---|---|---|---|
| **Armed conflict** | Armed conflict during the years 2020-2021 that caused at least 100 battle-related deaths | 2020-2021 | Battle-Related Deaths Dataset (version 22.1) (Pettersson et al. 2021) |
| **Tax revenue (% GDP) (ln)** | Total tax revenue as a percentage of GDP. | 2019 | Heritage Foundation (n.d.) |
| **Impartial public administration** | Measures extent to which public officials perform their duties in a rigorous and impartial manner. | 2019 | Varieties of Democracy (V-Dem) Project (Coppedge and et al 2023) (variable name: v2clrspct) |
| **Mean war mortality (ln)** | Mean war mortality rate for the years 2010-2019 | 2019 | GBD Collaborative Network (2020a) (variable name: mx_warterror_10years) |
| **Democracy-Dictatorship Index** | Dichotomous measure of democracy. A country is defined as democratic, if elections were conducted, these were free and fair, and if there was a peaceful turnover of legislative and executive offices following those elections. | 2019 | Bjørnskov and Rode (2020) (variable name: Democracy). This index is an update of Cheibub et al (2009). |
| **Lexical Index of Electoral Democracy** | Incorporates binary coding of different features of political regimes, which are aggregated together using the cumulative logic of a lexical scale. | 2019 | Skaaning, Gerring, and Bartusevičius (2015) (variable name: lexical_index) |
| **Polity2 Index** | Polychotomous measure of democracy. Captures the extent to which political leaders are selected via competitive elections and the extent to which there are constraints on the exercise of power by the executive. | 2018 | Marshall and Gurr (2020) (variable name: polity2) |
| **Machine Learning (ML) Democracy Index** | Continuous measure of democracy that captures political competition, political participation, and freedom of opinion. A machine learning approach was used to aggregate 10 regime characteristics (7 objective and 3 subjective/expert-based). | 2019 | Gründler and Krieger (2021) (variable name: c_mldi) |
| **Undercount ratio (ln) (WHO)** | Ratio of cumulative excess deaths due to COVID-19 (WHO estimates) over cumulative reported deaths due to COVID-19 | January 2020-December 31 2021 | WHO (2022) and Dong et al (2020) |
| **Undercount ratio (ln) (The Economist)** | Ratio of cumulative excess deaths due to COVID-19 (The Economist estimates) over cumulative reported deaths due to COVID-19 | January 2020-December 27 2021 | The Economist ([2020] 2022) and Dong et al (2020) |
| **Undercount ratio (ln) (Karlinsky & Kobak)** | Ratio of cumulative excess deaths due to COVID-19 (Karlinsky & Kobak estimates) over cumulative reported deaths due to COVID-19 | January 2020-December 31 2021 | Karlinsky and Kobak (2021) and Dong et al (2020) |

### 2. Summary statistics

| Variable | N | Missing (%) | Mean | SD | P25 | Median | P75 | Min | Max |
|---|---|---|---|---|---|---|---|---|---|
| Undercount ratio (IHME) (ln) | 185 | 13.15 | 2.31 | 1.61 | 1.22 | 1.87 | 3.61 | -3.53 | 6.15 |
| Kolmogorov-Smirnov statistic (ln) | 183 | 14.08 | -2.77 | 0.56 | -3.06 | -2.76 | -2.4 | -4.32 | -1.45 |
| Underdispersion index (ln) | 201 | 5.63 | -0.18 | 0.75 | -0.37 | -0.12 | 0 | -3.22 | 2.76 |
| Electoral democracy index | 172 | 19.25 | 0.53 | 0.25 | 0.3 | 0.53 | 0.76 | 0.02 | 0.92 |
| GDP per capita (ln) | 192 | 9.86 | 9.25 | 1.28 | 8.26 | 9.37 | 10.26 | 4.88 | 12.66 |
| Health service capacity & access | 184 | 13.62 | 64.43 | 29.33 | 34.4 | 76.09 | 90.65 | 10.77 | 99.67 |
| Information-gathering capacity | 174 | 18.31 | 0.02 | 0.11 | -0.04 | 0.02 | 0.09 | -0.31 | 0.27 |
| Prevalence of non-communicable diseases | 192 | 9.86 | 0.96 | 0.03 | 0.93 | 0.96 | 0.98 | 0.9 | 0.99 |
| Prevalence of lower respiratory diseases | 192 | 9.86 | 0 | 0 | 0 | 0 | 0 | 0 | 0 |
| Median age | 188 | 11.74 | 29.16 | 8.82 | 21.2 | 28.3 | 37.65 | 14.9 | 46.4 |
| Latin America & Caribbean | 213 | 0 | 0.21 | 0.41 | 0 | 0 | 0 | 0 | 1 |
| South Asia | 213 | 0 | 0.04 | 0.19 | 0 | 0 | 0 | 0 | 1 |
| Sub-Saharan Africa | 213 | 0 | 0.23 | 0.42 | 0 | 0 | 0 | 0 | 1 |
| Europe & Central Asia | 213 | 0 | 0.28 | 0.45 | 0 | 0 | 1 | 0 | 1 |
| East Asia & Pacific | 213 | 0 | 0.14 | 0.34 | 0 | 0 | 0 | 0 | 1 |
| Middle East & North Africa | 213 | 0 | 0.1 | 0.3 | 0 | 0 | 0 | 0 | 1 |
| North America | 213 | 0 | 0.01 | 0.12 | 0 | 0 | 0 | 0 | 1 |
| Island state | 213 | 0 | 0.3 | 0.46 | 0 | 0 | 1 | 0 | 1 |
| Disaster (2020-2021) | 213 | 0 | 0.38 | 0.49 | 0 | 0 | 1 | 0 | 1 |
| Armed conflict (2020-2021) | 213 | 0 | 0.13 | 0.34 | 0 | 0 | 0 | 0 | 1 |
| Tax revenue (% GDP) (ln) | 184 | 13.62 | 3.4 | 1.07 | 3.26 | 3.69 | 4.06 | 0 | 4.53 |
| Impartial public administration | 172 | 19.25 | 0.45 | 1.46 | -0.58 | 0.41 | 1.29 | -2.58 | 4.05 |
| Mean war mortality (2010-2019) (ln) | 192 | 9.86 | 0 | 0 | 0 | 0 | 0 | 0 | 0 |
| Democracy-Dictatorship index | 197 | 7.51 | 0.64 | 0.48 | 0 | 1 | 1 | 0 | 1 |
| Lexical Index of Electoral Democracy | 188 | 11.74 | 4.62 | 2.01 | 3 | 6 | 6 | 0 | 6 |
| Polity2 index | 163 | 23.47 | 4.23 | 6.07 | -1 | 7 | 9 | -10 | 10 |
| Continuous ML Democracy Index | 180 | 15.49 | 0.69 | 0.34 | 0.41 | 0.86 | 0.96 | 0 | 1 |
| Undercount ratio (WHO) (ln) | 181 | 15.02 | 1.85 | 2.11 | 1.04 | 1.79 | 2.96 | -5.59 | 7.38 |
| Undercount ratio (Economist) (ln) | 197 | 7.51 | 1.87 | 1.64 | 1.02 | 1.45 | 2.91 | -5.56 | 6.21 |
| Undercount ratio (Karlinksy & Kobak) (ln) | 100 | 53.05 | 0.95 | 1.3 | 0.6 | 1.02 | 1.4 | -4.69 | 5.06 |

### 3. Variable correlation matrix

| | Undercount ratio (IHME) (ln) | Kolmogorov-Smirnov statistic (ln) | Under-dispersion index (ln) | Electoral democracy index | GDP per capita (ln) | Health service capacity & access | Information-gathering capacity | Prevalence of non-communicable diseases | Prevalence of lower respiratory diseases | Median age | Island state | Disaster (2020-2021) | Armed conflict (2020-2021) | Tax revenue (% GDP) (ln) | Impartial public administration | Mean war mortality (2010-2019) (ln) | Democracy-Dictatorship index | Lexical Index of Electoral Democracy | Polity2 index | Continuous ML Democracy Index | Undercount ratio (WHO) (ln) | Undercount ratio (Economist) (ln) | Undercount ratio (Karlinksy & Kobak) (ln) |
|---|---|---|---|---|---|---|---|---|---|---|---|---|---|---|---|---|---|---|---|---|---|---|---|
| Undercount ratio (IHME) (ln) | 1 | | | | | | | | | | | | | | | | | | | | | | |
| Kolmogorov-Smirnov statistic (ln) | 0.170 | 1 | | | | | | | | | | | | | | | | | | | | | |
| Underdispersion index (ln) | 0.316*** | 0.391*** | 1 | | | | | | | | | | | | | | | | | | | | |
| Electoral democracy index | -0.471*** | -0.395*** | -0.387*** | 1 | | | | | | | | | | | | | | | | | | | |
| GDP per capita (ln) | -0.562*** | -0.231** | -0.276** | 0.478*** | 1 | | | | | | | | | | | | | | | | | | |
| Health service capacity & access | -0.258** | -0.258** | -0.113 | 0.384*** | 0.523*** | 1 | | | | | | | | | | | | | | | | | |
| Information-gathering capacity | -0.164 | -0.273** | -0.131 | 0.320*** | 0.249** | 0.0693 | 1 | | | | | | | | | | | | | | | | |
| Prevalence of non-communicable diseases | -0.417*** | -0.398*** | -0.149 | 0.450*** | 0.768*** | 0.665*** | 0.171 | 1 | | | | | | | | | | | | | | | |
| Prevalence of lower respiratory diseases | 0.221* | 0.124 | -0.127 | -0.186 | -0.551*** | -0.389*** | -0.132 | -0.498*** | 1 | | | | | | | | | | | | | | |
| Median age | -0.381*** | -0.380*** | -0.207* | 0.559*** | 0.675*** | 0.643*** | 0.301*** | 0.858*** | -0.348*** | 1 | | | | | | | | | | | | | |
| Island state | -0.389*** | 0.173 | -0.117 | 0.0816 | 0.105 | -0.0867 | -0.0977 | -0.0609 | -0.0574 | 0.0453 | 1 | | | | | | | | | | | | |
| Disaster (2020-2021) | 0.221* | 0.0422 | -0.0707 | -0.0342 | -0.225* | -0.146 | -0.200* | -0.199* | 0.00681 | -0.187 | -0.0709 | 1 | | | | | | | | | | | |
| Armed conflict (2020-2021) | 0.133 | -0.0563 | 0.0586 | -0.152 | -0.169 | -0.103 | 0.0596 | -0.122 | 0.0949 | -0.148 | -0.00170 | -0.141 | 1 | | | | | | | | | | |
| Tax revenue (% GDP) (ln) | -0.106 | -0.337*** | -0.206* | 0.582*** | 0.167 | 0.511*** | 0.170 | 0.404*** | -0.134 | 0.565*** | -0.0401 | 0.0312 | -0.0344 | 1 | | | | | | | | | |
| Impartial public administration | -0.567*** | -0.277** | -0.255** | 0.768*** | 0.724*** | 0.497*** | 0.272** | 0.607*** | -0.430*** | 0.608*** | 0.161 | -0.150 | -0.207* | 0.416*** | 1 | | | | | | | | |
| Mean war mortality (2010-2019) (ln) | 0.0953 | -0.132 | 0.177 | -0.239** | -0.232** | -0.0859 | -0.184 | -0.0937 | 0.0739 | -0.273** | -0.100 | 0.0850 | 0.444*** | -0.0981 | -0.285** | 1 | | | | | | | |
| Democracy-Dictatorship index | -0.369*** | -0.402*** | -0.342*** | 0.825*** | 0.226** | 0.165 | 0.367*** | 0.309*** | -0.157 | 0.453*** | 0.0413 | 0.00260 | -0.0474 | 0.548*** | 0.542*** | -0.222* | 1 | | | | | | |
| Lexical Index of Electoral Democracy | -0.339*** | -0.383*** | -0.348*** | 0.847*** | 0.216* | 0.261** | 0.301*** | 0.258** | -0.0753 | 0.423*** | 0.0199 | 0.0295 | -0.0112 | 0.604*** | 0.481*** | -0.0329 | 0.839*** | 1 | | | | | |
| Polity2 index | -0.306*** | -0.359*** | -0.288** | 0.855*** | 0.196* | 0.188 | 0.354*** | 0.247** | -0.115 | 0.422*** | 0.0658 | 0.124 | -0.157 | 0.658*** | 0.521*** | -0.146 | 0.866*** | 0.896*** | 1 | | | | |
| Continuous ML Democracy Index | -0.399*** | -0.454*** | -0.384*** | 0.875*** | 0.260** | 0.270** | 0.320*** | 0.317*** | -0.126 | 0.449*** | -0.00668 | 0.0695 | -0.137 | 0.626*** | 0.549*** | -0.0860 | 0.895*** | 0.923*** | 0.927*** | 1 | | | |
| Undercount ratio (WHO) (ln) | 0.746*** | 0.238** | 0.277** | -0.534*** | -0.458*** | -0.270** | -0.174 | -0.403*** | 0.226** | -0.321*** | -0.217* | 0.154 | 0.168 | -0.216* | -0.531*** | 0.0763 | -0.436*** | -0.382*** | -0.400*** | -0.463*** | 1 | | |
| Undercount ratio (Economist) (ln) | 0.854*** | 0.292** | 0.330*** | -0.586*** | -0.531*** | -0.270** | -0.180 | -0.424*** | 0.218* | -0.380*** | -0.236** | 0.136 | 0.145 | -0.193* | -0.622*** | 0.0751 | -0.496*** | -0.437*** | -0.428*** | -0.525*** | 0.861*** | 1 | |
| Undercount ratio (Karlinksy & Kobak) (ln) | 0.792*** | 0.324*** | 0.312*** | -0.589*** | -0.511*** | -0.205* | -0.191* | -0.397*** | 0.219* | -0.349*** | -0.262** | 0.175 | 0.164 | -0.142 | -0.599*** | 0.0922 | -0.482*** | -0.422*** | -0.421*** | -0.498*** | 0.878*** | 0.965*** | 1 |

* p<0.1; ** p<0.05; *** p<0.01

### 4. Coefficient correlation matrices

#### 4.1 Undercount ratio

| | Electoral democracy index | GDP per capita (ln) | Health service capacity & access | Information-gathering capacity | Prevalence of non-communicable diseases | Prevalence of lower respiratory diseases | Median age | Latin America & Caribbean | South Asia | Sub-Saharan Africa | Europe & Central Asia | East Asia & Pacific | Middle East & North Africa | Constant |
|---|---|---|---|---|---|---|---|---|---|---|---|---|---|---|
| Electoral democracy index | 1.000 | | | | | | | | | | | | | |
| GDP per capita (ln) | 0.068 | 1.000 | | | | | | | | | | | | |
| Health service capacity & access | -0.000 | -0.443 | | | | | | | | | | | | |
| Information-gathering capacity | -0.267 | -0.175 | -0.147 | 1.000 | | | | | | | | | | |
| Prevalence of non-communicable diseases | 0.187 | -0.071 | -0.210 | -0.039 | 1.000 | | | | | | | | | |
| Prevalence of lower respiratory diseases | 0.032 | 0.320 | -0.224 | -0.079 | 0.044 | 1.000 | | | | | | | | |
| Median age | -0.197 | -0.309 | -0.222 | 0.149 | -0.467 | -0.152 | 1.000 | | | | | | | |
| Latin America & Caribbean | 0.127 | 0.004 | -0.093 | -0.235 | 0.094 | -0.539 | 0.370 | 1.000 | | | | | | |
| South Asia | 0.365 | -0.102 | 0.171 | -0.172 | 0.241 | -0.418 | 0.140 | 0.581 | 1.000 | | | | | |
| Sub-Saharan Africa | 0.306 | -0.215 | 0.136 | -0.061 | 0.318 | -0.424 | 0.303 | 0.683 | 0.773 | 1.000 | | | | |
| Europe & Central Asia | 0.459 | 0.355 | 0.220 | -0.464 | 0.236 | -0.092 | -0.588 | 0.123 | 0.253 | 0.128 | 1.000 | | | |
| East Asia & Pacific | 0.513 | -0.201 | 0.181 | -0.186 | 0.283 | -0.412 | 0.164 | 0.640 | 0.679 | 0.826 | 0.273 | 1.000 | | |
| Middle East & North Africa | 0.577 | -0.161 | 0.005 | -0.090 | -0.077 | -0.278 | 0.500 | 0.547 | 0.608 | 0.709 | 0.051 | 0.719 | 1.000 | |
| Constant | -0.225 | -0.029 | 0.265 | 0.073 | -0.990 | -0.086 | 0.439 | -0.143 | -0.281 | -0.364 | -0.264 | -0.320 | 0.019 | 1.000 |

*Note*: Based on the baseline OLS regression (column 2 of Table 1 in the main text)

#### 4.2 Benford-noncompliance

| | Electoral democracy index | GDP per capita (ln) | Health service capacity & access | Information-gathering capacity | Prevalence of non-communicable diseases | Prevalence of lower respiratory diseases | Median age | Latin America & Caribbean | South Asia | Sub-Saharan Africa | Europe & Central Asia | East Asia & Pacific | Middle East & North Africa | Magnitude<1000 | Constant |
|---|---|---|---|---|---|---|---|---|---|---|---|---|---|---|---|
| Electoral democracy index | 1.000 | | | | | | | | | | | | | | |
| GDP per capita (ln) | -0.098 | 1.000 | | | | | | | | | | | | | |
| Health service capacity & access | 0.168 | -0.123 | 1.000 | | | | | | | | | | | | |
| Information-gathering capacity | -0.349 | 0.195 | -0.107 | 1.000 | | | | | | | | | | | |
| Prevalence of non-communicable diseases | -0.192 | -0.165 | -0.255 | -0.034 | 1.000 | | | | | | | | | | |
| Prevalence of lower respiratory diseases | -0.141 | 0.449 | 0.106 | -0.017 | -0.103 | 1.000 | | | | | | | | | |
| Median age | -0.174 | -0.281 | -0.282 | 0.026 | -0.439 | -0.073 | 1.000 | | | | | | | | |
| Latin America & Caribbean | 0.052 | -0.039 | 0.004 | -0.060 | 0.055 | -0.095 | 0.088 | 1.000 | | | | | | | |
| South Asia | 0.111 | -0.094 | 0.120 | -0.197 | 0.207 | -0.175 | -0.070 | 0.860 | 1.000 | | | | | | |
| Sub-Saharan Africa | 0.037 | -0.041 | 0.174 | 0.007 | 0.065 | -0.100 | 0.102 | 0.931 | 0.867 | 1.000 | | | | | |
| Europe & Central Asia | 0.088 | 0.039 | 0.037 | -0.092 | 0.003 | -0.025 | -0.054 | 0.938 | 0.840 | 0.882 | 1.000 | | | | |
| East Asia & Pacific | 0.176 | 0.011 | 0.147 | -0.146 | 0.005 | -0.021 | -0.044 | 0.921 | 0.846 | 0.904 | 0.940 | 1.000 | | | |
| Middle East & North Africa | 0.282 | -0.057 | 0.094 | -0.071 | -0.152 | -0.104 | 0.139 | 0.903 | 0.791 | 0.889 | 0.896 | 0.897 | 1.000 | | |
| Magnitude<1000 | -0.194 | 0.218 | 0.022 | 0.401 | 0.089 | 0.067 | 0.041 | 0.021 | -0.023 | 0.097 | -0.033 | -0.015 | -0.030 | 1.000 | |
| Constant | 0.200 | 0.040 | 0.248 | 0.022 | -0.982 | 0.008 | 0.429 | -0.164 | -0.297 | -0.185 | -0.111 | -0.117 | 0.049 | -0.142 | 1.000 |

*Note*: Based on the baseline OLS regression (column 4 of Table 1 in the main text)

#### 4.3 Underdispersion index

| | Electoral democracy index | GDP per capita (ln) | Health service capacity & access | Information-gathering capacity | Prevalence of non-communicable diseases | Prevalence of lower respiratory diseases | Median age | Latin America & Caribbean | South Asia | Sub-Saharan Africa | Europe & Central Asia | East Asia & Pacific | Middle East & North Africa | Constant |
|---|---|---|---|---|---|---|---|---|---|---|---|---|---|---|
| Electoral democracy index | 1.000 | | | | | | | | | | | | | |
| GDP per capita (ln) | 0.233 | 1.000 | | | | | | | | | | | | |
| Health service capacity & access | -0.009 | -0.184 | 1.000 | | | | | | | | | | | |
| Information-gathering capacity | -0.363 | -0.015 | -0.061 | 1.000 | | | | | | | | | | |
| Prevalence of non-communicable diseases | -0.105 | -0.250 | -0.135 | -0.022 | 1.000 | | | | | | | | | |
| Prevalence of lower respiratory diseases | 0.054 | 0.143 | 0.453 | 0.131 | 0.136 | 1.000 | | | | | | | | |
| Median age | -0.123 | -0.346 | -0.315 | -0.123 | -0.450 | -0.303 | 1.000 | | | | | | | |
| Latin America & Caribbean | 0.058 | -0.016 | 0.069 | -0.110 | 0.009 | -0.001 | 0.030 | | | | | | | |
| South Asia | 0.100 | -0.013 | 0.045 | -0.120 | 0.092 | -0.005 | -0.005 | 0.955 | 1.000 | | | | | |
| Sub-Saharan Africa | 0.089 | -0.036 | 0.126 | -0.098 | 0.053 | 0.020 | 0.020 | 0.958 | 0.980 | 1.000 | | | | |
| Europe & Central Asia | 0.051 | 0.056 | 0.019 | -0.111 | -0.007 | -0.006 | -0.007 | 0.955 | 0.960 | 0.956 | 1.000 | | | |
| East Asia & Pacific | 0.105 | 0.020 | 0.009 | -0.086 | 0.058 | 0.013 | -0.015 | 0.960 | 0.973 | 0.972 | 0.973 | 1.000 | | |
| Middle East & North Africa | 0.171 | 0.003 | 0.012 | -0.116 | -0.052 | -0.004 | 0.080 | 0.947 | 0.955 | 0.958 | 0.957 | 0.966 | 1.000 | |
| Constant | 0.041 | 0.149 | 0.112 | 0.060 | -0.978 | -0.212 | 0.467 | -0.163 | -0.250 | -0.213 | -0.151 | -0.217 | -0.107 | 1.000 |

*Note*: Based on the baseline OLS regression (column 7 of Table 1 in the main text)

### 5. Complete results

#### 5.1 Democracy and undercounting

| | (1) | (2) | (3) | (4) | (5) |
|---|---|---|---|---|---|
| Dependent variable | Undercount ratio (IHME) (ln) | Undercount ratio (IHME) (ln) | Undercount ratio (IHME) (ln) | Undercount ratio (IHME) (ln) | Undercount ratio (IHME) (ln) |
| Electoral Democracy Index | -3.398*** | -2.996*** | -1.545*** | -1.487*** | -1.282*** |
| | (0.435) | (0.504) | (0.403) | (0.412) | (0.387) |
| GDP per capita (ln) | | | -0.527*** | -0.530*** | -0.386*** |
| | | | (0.122) | (0.123) | (0.127) |
| Health service capacity & access | | | -0.0188*** | -0.0182*** | -0.0104* |
| | | | (0.00569) | (0.00574) | (0.00618) |
| Information-gathering capacity | | | | -0.284 | -0.241 |
| | | | | (0.863) | (0.783) |
| Prevalence of non-communicable diseases | | | | | -25.50*** |
| | | | | | (8.540) |
| Prevalence of lower respiratory diseases | | | | | 42.51 |
| | | | | | (194.4) |
| Median age | | | | | 0.00652 |
| | | | | | (0.0295) |
| Latin America & Caribbean | | 0.208 | -0.794*** | -0.760*** | -0.980*** |
| | | (0.180) | (0.201) | (0.223) | (0.280) |
| South Asia | | 0.649 | -0.943** | -0.899* | -1.304*** |
| | | (0.469) | (0.453) | (0.467) | (0.475) |
| Sub-Saharan Africa | | 1.604*** | -0.675** | -0.615* | -0.967** |
| | | (0.271) | (0.341) | (0.350) | (0.444) |
| Europe & Central Asia | | -0.0502 | -0.371** | -0.338* | -0.264 |
| | | (0.159) | (0.152) | (0.185) | (0.206) |
| East Asia & Pacific | | -0.744 | -1.475*** | -1.436*** | -1.701*** |
| | | (0.524) | (0.463) | (0.482) | (0.513) |
| Middle East & North Africa | | -0.536 | -0.800*** | -0.765*** | -0.624** |
| | | (0.324) | (0.264) | (0.268) | (0.292) |
| Constant | 4.179*** | 3.622*** | 9.963*** | 9.892*** | 32.19*** |
| | (0.263) | (0.422) | (1.059) | (1.105) | (7.781) |
| Adjusted R-squared | 0.270 | 0.524 | 0.696 | 0.696 | 0.709 |
| Countries | 171 | 171 | 169 | 168 | 168 |

*Note*: Undercount ratio as of December 31 2021, based on excess mortality estimated by IHME. Reference category for region is North America. Robust standard errors are reported in parenthesis. *p<0.1; **p<0.05; ***p<0.01

#### 5.2 Democracy and Benford-noncompliance

| | (1) | (2) | (3) | (4) | (5) | (6) |
|---|---|---|---|---|---|---|
| Dependent variable | Kolmogorov-Smirnov statistic (ln) | Kolmogorov-Smirnov statistic (ln) | Kolmogorov-Smirnov statistic (ln) | Kolmogorov-Smirnov statistic (ln) | Kolmogorov-Smirnov statistic (ln) | Kolmogorov-Smirnov statistic (ln) |
| Electoral Democracy Index | -0.646*** | -0.534*** | -0.749*** | -0.774*** | -0.666*** | -0.662** |
| | (0.161) | (0.203) | (0.211) | (0.225) | (0.242) | (0.274) |
| GDP per capita (ln) | | | 0.132** | 0.135** | 0.174** | 0.186** |
| | | | (0.0646) | (0.0628) | (0.0722) | (0.0784) |
| Health service capacity & access | | | -0.000583 | -0.000919 | 0.00183 | 0.00225 |
| | | | (0.00329) | (0.00332) | (0.00378) | (0.00440) |
| Information-gathering capacity | | | | 0.00727 | 0.0519 | -0.379 |
| | | | | (0.561) | (0.548) | (0.616) |
| Prevalence of non-communicable diseases | | | | | -3.034 | -5.141 |
| | | | | | (4.740) | (5.513) |
| Prevalence of lower respiratory diseases | | | | | -54.55 | -26.59 |
| | | | | | (149.8) | (156.9) |
| Median age | | | | | -0.0162 | -0.00679 |
| | | | | | (0.0158) | (0.0177) |
| Latin America & Caribbean | | 0.562 | 0.697 | 0.692 | 0.620 | 0.663 |
| | | (0.410) | (0.434) | (0.435) | (0.457) | (0.466) |
| South Asia | | 0.732 | 0.905* | 0.888* | 0.817 | 0.855 |
| | | (0.470) | (0.494) | (0.504) | (0.528) | (0.559) |
| Sub-Saharan Africa | | 0.443 | 0.672 | 0.641 | 0.540 | 0.629 |
| | | (0.410) | (0.470) | (0.471) | (0.491) | (0.503) |
| Europe & Central Asia | | 0.427 | 0.484 | 0.480 | 0.535 | 0.554 |
| | | (0.404) | (0.423) | (0.427) | (0.442) | (0.452) |
| East Asia & Pacific | | 0.413 | 0.470 | 0.459 | 0.424 | 0.508 |
| | | (0.422) | (0.450) | (0.457) | (0.467) | (0.485) |
| Middle East & North Africa | | 0.419 | 0.448 | 0.433 | 0.400 | 0.502 |
| | | (0.437) | (0.456) | (0.459) | (0.485) | (0.506) |
| Magnitude<1000 | | 0.404*** | 0.480*** | 0.459*** | 0.455*** | |
| | | (0.102) | (0.115) | (0.126) | (0.122) | |
| Constant | -2.461*** | -3.058*** | -4.249*** | -4.227*** | -1.342 | 0.174 |
| | (0.0918) | (0.430) | (0.728) | (0.706) | (4.194) | (4.768) |
| Adjusted R-squared | 0.080 | 0.151 | 0.171 | 0.162 | 0.164 | 0.075 |
| Observations | 169 | 169 | 167 | 166 | 166 | 135 |

Note: Kolmogorov-Smirnov test statistic as of December 31 2021. Reference category for region is North America. Magnitude<1000 is a dummy variable identifying those countries without at least 1000 cases or deaths on any single day. Column 6 excludes those countries from the sample. Robust standard errors are reported in parenthesis. *p<0.1; **p<0.05; ***p<0.01

##### 5.3 Democracy and underdispersion

| | (1) | (2) | (3) | (4) | (5) |
|---|---|---|---|---|---|
| Dependent variable | Underdispersion index (ln) | Underdispersion index (ln) | Underdispersion index (ln) | Underdispersion index (ln) | Underdispersion index (ln) |
| Electoral Democracy Index | -1.080*** | -1.158*** | -1.079*** | -1.118*** | -1.031*** |
| | (0.251) | (0.307) | (0.300) | (0.318) | (0.330) |
| GDP per capita (ln) | | | -0.200** | -0.205*** | -0.210** |
| | | | (0.0776) | (0.0778) | (0.0856) |
| Health service capacity & access | | | 0.00984** | 0.00941* | 0.0110** |
| | | | (0.00469) | (0.00481) | (0.00521) |
| Information-gathering capacity | | | | 0.375 | 0.441 |
| | | | | (0.781) | (0.780) |
| Prevalence of non-communicable diseases | | | | | 0.577 |
| | | | | | (6.008) |
| Prevalence of lower respiratory diseases | | | | | -259.7 |
| | | | | | (204.2) |
| Median age | | | | | -0.0179 |
| | | | | | (0.0182) |
| Latin America & Caribbean | | 1.577* | 1.531* | 1.490* | 1.516* |
| | | (0.852) | (0.837) | (0.835) | (0.834) |
| South Asia | | 1.382 | 1.472* | 1.431* | 1.491* |
| | | (0.848) | (0.834) | (0.833) | (0.833) |
| Sub-Saharan Africa | | 1.314 | 1.453* | 1.413* | 1.423* |
| | | (0.835) | (0.839) | (0.837) | (0.836) |
| Europe & Central Asia | | 1.823** | 1.773** | 1.733** | 1.788** |
| | | (0.835) | (0.814) | (0.812) | (0.815) |
| East Asia & Pacific | | 1.531* | 1.578* | 1.540* | 1.569* |
| | | (0.841) | (0.819) | (0.815) | (0.816) |
| Middle East & North Africa | | 1.844** | 1.916** | 1.891** | 1.860** |
| | | (0.851) | (0.829) | (0.825) | (0.831) |
| Constant | 0.379*** | -1.146 | -0.0229 | 0.0982 | 0.283 |
| | (0.143) | (0.862) | (1.013) | (1.037) | (5.396) |
| Adjusted R-squared | 0.105 | 0.188 | 0.206 | 0.204 | 0.204 |
| Observations | 167 | 167 | 167 | 166 | 166 |

Note: Underdispersion index as of January 30 2022. Reference category for region is North America. Robust standard errors are reported in parenthesis.
*p<0.1; **p<0.05; ***p<0.01

#### 6. Shapley decomposition for covariate groups

Figure 1 presents the Shapley decomposition of the R-squared for each of the dependent variables, based on the covariate groups described in the methods section. In the case of undercounting, for example, the share of the explained variance captured by democracy is 11.57%, while for health system capacity, information-gathering capacity, and pandemic vulnerability it is 31.39 %, 8.76%, and 26.89% respectively. The two measures of statistical irregularity – Benford-noncompliance and underdispersion - are less likely to reflect unintentional mismeasurement. Unsurprisingly, therefore, the three covariate groups designed to control for unintended misreporting capture less of the explained variation in those two dependent variables.

**Figure 1: Shapley decomposition by covariate group**

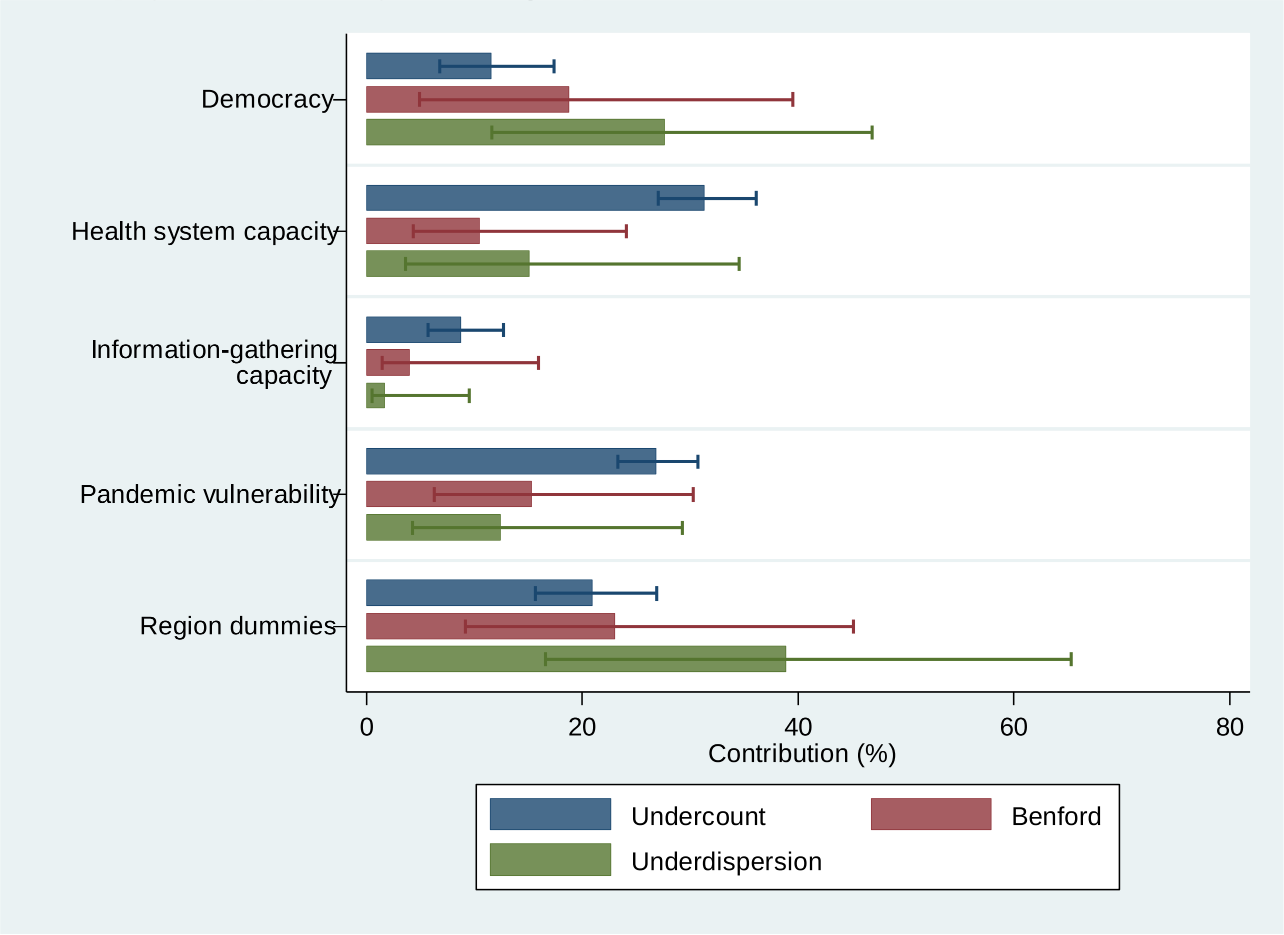

*Note*: The legend indicates the dependent variables and the horizontal bars represent the percentage of the R-squared that is captured by each covariate group. 95% confidence intervals calculated based on 2000 bootstrap replications.

#### 7. First digit analysis of reported cases and deaths

The results presented in columns 3-5 in Table 1 in the main text are based on a dependent variable that reflects the extent to which reported cases and deaths fail to conform with Benford's Law. That law posits that the first digits in non-manipulated data should accord with a distribution where the number 1 is the most likely to occur and the remaining digits are increasingly less likely to occur. Deviation from this pattern can be used to estimate whether reported data are subject to manipulation (Tam Cho and Gaines, 2012). Along with the underdispersion index, this provides a more direct way to examine the relationship between regime-type and manipulation. However, there remains the possibility that adroit autocratic leaders may arrange the digits so they accord with Benford's Law (Diekmann 2007; Michalski and Stoltz 2013).

To perform the digit analysis I used the reported daily cases and deaths recorded by the Johns Hopkins Center for Systems Science and Engineering for the period 22 January 2020 to 31 December 2021 (Dong, Du, and Gardner 2020). Tests for Benfordness require a sufficient number of observations and numerical values that are spread over several orders of magnitude (Fewster 2009; Goodman 2016). For the baseline results I, therefore, I exclude those countries with 100 or less observations and include a dummy variable identifying those countries with no day in which at least 1,000 cases or deaths were reported (main text Table 1, column 4). I also run the regression without those countries that do not fulfil the latter condition (main text Table 1, column 5).

In order to measure Benford-noncompliance I used the Kolmogorov-Smirnov test statistic, which measures the extent to which the first digit frequencies deviate from those that would be expected based on Benford's Law. That test statistic was produced using the DIGDIS module in Stata (version 14.1). Exact p-values were estimated using

Monte Carlo simulation (with 10,000 replications). In the table below I report the Kolmogorov-Smirnov statistic and p-values, along with three further tests for Benford-noncompliance (Pearson's $X^2$, likelihood ratio, and Cressie–Read's $D^2$). Larger test statistics that are statistically significant indicate deviation from Benford's law and, therefore, the potential presence of manipulation. The last column flags those countries where there was no day with at least 1,000 reported cases or deaths. The Kolmogorov-Smirnov statistic is more appropriate in the context of an epidemic because it is better equipped to handle smaller sample sizes. The results for that statistic indicate that (for those countries that meet the magnitude criterion) the reported data for the Maldives is the least Benford-compliant and the reported data for Israel is the most Benford-compliant. The mean for that statistic (again for those countries that fulfill the magnitude criterion) is 0.063 (median 0.054; inter-quartile range 0.041-0.076).

**Goodness-of-fit tests**

| Country | iso3 | Obs | Kolmogorov-Smirnov | p-value | Pearson's $X^2$ | p-value | Likelihood ratio | p-value | Cressie–Read's D2 | p-value | No daily cases or deaths ≥ 1000 |
|---|---|---|---|---|---|---|---|---|---|---|---|
| Brunei | BRN | 290 | 0.2334528 | 0 | 79.84755 | 0 | 75.448 | 0 | 77.85403 | 0 | 1 |
| Maldives | MDV | 794 | 0.2304562 | 0 | 206.9632 | 0 | 191.9025 | 0 | 200.7339 | 0 | 0 |
| Nicaragua | NIC | 191 | 0.212059 | 0 | 51.25778 | 0 | 52.18241 | 0 | 51.1476 | 0 | 1 |
| Belarus | BLR | 1285 | 0.206363 | 0 | 694.2657 | 0 | 699.5508 | 0 | 681.2028 | 0 | 0 |
| Qatar | QAT | 934 | 0.1973969 | 0 | 153.9572 | 0 | 161.4358 | 0 | 155.6976 | 0 | 0 |
| Chad | TCD | 545 | 0.1870989 | 0 | 86.45919 | 0 | 89.19609 | 0 | 86.93574 | 0 | 1 |
| Tajikistan | TJK | 423 | 0.1792489 | 0 | 207.2901 | 0 | 171.3788 | 0 | 191.5454 | 0 | 1 |
| Sao Tome and Principe | STP | 425 | 0.1487611 | 0 | 49.46038 | 0 | 52.79668 | 0 | 50.29157 | 0 | 1 |
| Barbados | BRB | 596 | 0.14794 | 0 | 71.73673 | 0 | 79.22414 | 0 | 73.7731 | 0 | 1 |
| Burkina Faso | BFA | 612 | 0.146306 | 0 | 74.92445 | 0 | 79.17616 | 0 | 75.91194 | 0 | 1 |
| Eritrea | ERI | 404 | 0.1441659 | 0 | 37.03986 | 0 | 38.14818 | 0 | 37.31694 | 0 | 1 |
| Togo | TGO | 754 | 0.1422421 | 0 | 69.18964 | 0 | 70.79977 | 0 | 69.53342 | 0 | 1 |
| Antigua and Barbuda | ATG | 302 | 0.1396619 | 0 | 36.9121 | 0.0001 | 40.07162 | 0 | 37.74008 | 0.0001 | 1 |
| Bhutan | BTN | 349 | 0.1389246 | 0 | 42.55801 | 0 | 46.3595 | 0 | 43.4396 | 0 | 1 |
| South Sudan | SSD | 432 | 0.1386195 | 0 | 45.93952 | 0 | 46.14129 | 0 | 45.86076 | 0 | 1 |
| Turkey | TUR | 1311 | 0.1384394 | 0 | 181.6705 | 0 | 193.016 | 0 | 184.257 | 0 | 0 |
| Australia | AUS | 984 | 0.1359619 | 0 | 100.1658 | 0 | 97.39376 | 0 | 98.98324 | 0 | 0 |
| Gabon | GAB | 435 | 0.1355128 | 0 | 41.44035 | 0 | 48.14615 | 0 | 43.33617 | 0 | 0 |
| Iraq | IRQ | 1323 | 0.1339618 | 0 | 143.304 | 0 | 150.3823 | 0 | 145.1468 | 0 | 0 |
| Saint Kitts and Nevis | KNA | 218 | 0.1313052 | 0.0001 | 20.17817 | 0.0107 | 22.88218 | 0.0039 | 20.92677 | 0.0075 | 1 |
| Uzbekistan | UZB | 1069 | 0.1289065 | 0 | 131.7487 | 0 | 139.5176 | 0 | 133.7618 | 0 | 1 |
| Egypt | EGY | 1319 | 0.1251836 | 0 | 144.4836 | 0 | 155.1209 | 0 | 147.2117 | 0 | 0 |
| Costa Rica | CRI | 895 | 0.1246683 | 0 | 76.98255 | 0 | 75.50553 | 0 | 76.30042 | 0 | 0 |
| Timor-Leste | TLS | 371 | 0.1239569 | 0 | 34.21103 | 0 | 35.35822 | 0 | 34.49442 | 0 | 1 |
| Saint Lucia | LCA | 481 | 0.1216313 | 0 | 33.13869 | 0.0001 | 34.66374 | 0 | 33.55292 | 0.0001 | 1 |
| Liechtenstein | LIE | 517 | 0.121146 | 0 | 43.66111 | 0 | 51.32356 | 0 | 45.77078 | 0 | 1 |
| Malta | MLT | 868 | 0.1196529 | 0 | 58.05551 | 0 | 60.77365 | 0 | 58.78142 | 0 | 0 |
| Syrian Arab Republic | SYR | 1098 | 0.1157212 | 0 | 91.40367 | 0 | 89.85941 | 0 | 90.58826 | 0 | 1 |
| Mauritius | MUS | 384 | 0.1130325 | 0 | 31.77582 | 0.0002 | 32.51251 | 0.0002 | 31.91369 | 0.0002 | 0 |
| Mexico | MEX | 1265 | 0.1127319 | 0 | 118.5426 | 0 | 111.7813 | 0 | 115.9293 | 0 | 0 |
| Bangladesh | BGD | 1294 | 0.1089137 | 0 | 109.6735 | 0 | 109.4732 | 0 | 109.2163 | 0 | 0 |
| San Marino | SMR | 424 | 0.1076338 | 0 | 26.80256 | 0.0009 | 30.35322 | 0.0002 | 27.83646 | 0.0003 | 1 |
| Gambia | GMB | 405 | 0.106582 | 0 | 27.85495 | 0.0004 | 31.04191 | 0.0001 | 28.76198 | 0.0004 | 1 |
| Peru | PER | 1188 | 0.1059091 | 0 | 80.53629 | 0 | 77.82894 | 0 | 79.42559 | 0 | 0 |
| Saudi Arabia | SAU | 1219 | 0.1041473 | 0 | 127.156 | 0 | 136.0004 | 0 | 129.5164 | 0 | 0 |
| Guinea-Bissau | GNB | 382 | 0.1014128 | 0.0003 | 26.33275 | 0.0011 | 28.55627 | 0.0004 | 26.93342 | 0.0003 | 1 |
| Cuba | CUB | 1081 | 0.1000754 | 0 | 89.93485 | 0 | 83.99972 | 0 | 87.53856 | 0 | 0 |
| Sierra Leone | SLE | 543 | 0.099306 | 0 | 24.72649 | 0.0018 | 24.90876 | 0.0013 | 24.75637 | 0.0014 | 1 |
| Myanmar | MMR | 976 | 0.0977351 | 0 | 46.23128 | 0 | 48.706 | 0 | 46.96671 | 0 | 0 |
| Montenegro | MNE | 1092 | 0.0952531 | 0 | 100.022 | 0 | 93.33142 | 0 | 97.48561 | 0 | 0 |
| Azerbaijan | AZE | 1242 | 0.0952025 | 0 | 62.49203 | 0 | 64.42235 | 0 | 62.99819 | 0 | 0 |
| Dominica | DMA | 162 | 0.0940317 | 0.0379 | 10.03705 | 0.2612 | 10.57175 | 0.2384 | 10.17759 | 0.2521 | 1 |
| Nepal | NPL | 1146 | 0.0935594 | 0 | 51.63498 | 0 | 53.06041 | 0 | 52.04868 | 0 | 0 |
| Bahamas | BHS | 546 | 0.0924758 | 0 | 28.29237 | 0.0006 | 30.20065 | 0.0004 | 28.82927 | 0.0006 | 1 |
| United Arab Emirates | ARE | 1223 | 0.091317 | 0 | 53.06688 | 0 | 57.30115 | 0 | 54.31988 | 0 | 0 |
| Mozambique | MOZ | 982 | 0.091027 | 0 | 46.69217 | 0 | 46.26827 | 0 | 46.48189 | 0 | 0 |
| Laos | LAO | 381 | 0.0900461 | 0.0011 | 19.89649 | 0.0118 | 20.18593 | 0.0104 | 19.95247 | 0.0114 | 0 |
| Liberia | LBR | 371 | 0.0898056 | 0.0012 | 22.11693 | 0.005 | 23.03951 | 0.0034 | 22.36214 | 0.0043 | 1 |
| Czechia | CZE | 1242 | 0.0880961 | 0 | 55.51372 | 0 | 54.88775 | 0 | 55.23442 | 0 | 0 |

| | | | | | | | | | | | |
|---|---|---|---|---|---|---|---|---|---|---|---|
| Paraguay | PRY | 1128 | 0.0872679 | 0 | 73.57557 | 0 | 78.12093 | 0 | 74.866 | 0 | 0 |
| Cyprus | CYP | 911 | 0.0861947 | 0 | 35.59008 | 0.0001 | 37.91264 | 0.0001 | 36.28856 | 0.0001 | 0 |
| Taiwan | TWN | 650 | 0.0859557 | 0 | 28.8257 | 0.0005 | 31.6841 | 0.0002 | 29.62521 | 0.0003 | 1 |
| Equatorial Guinea | GNQ | 251 | 0.0854242 | 0.0169 | 20.89506 | 0.0092 | 23.97675 | 0.0023 | 21.74066 | 0.0064 | 0 |
| Congo | COG | 239 | 0.0841325 | 0.0209 | 10.98762 | 0.1978 | 12.44439 | 0.134 | 11.41246 | 0.1755 | 1 |
| Lithuania | LTU | 1146 | 0.0830882 | 0 | 36.85038 | 0.0001 | 37.92676 | 0.0001 | 37.16743 | 0.0001 | 0 |
| Mongolia | MNG | 771 | 0.082887 | 0 | 37.77774 | 0 | 38.31947 | 0 | 37.89907 | 0 | 0 |
| Grenada | GRD | 175 | 0.0828787 | 0.066 | 8.821613 | 0.357 | 9.502037 | 0.3143 | 9.026704 | 0.3419 | 1 |
| Fiji | FJI | 350 | 0.0818271 | 0.0065 | 16.22119 | 0.0417 | 16.28495 | 0.042 | 16.22031 | 0.041 | 0 |
| Central African Republic | CAF | 243 | 0.0816861 | 0.028 | 15.22034 | 0.0575 | 15.35676 | 0.0581 | 15.24175 | 0.0564 | 0 |
| Luxembourg | LUX | 769 | 0.0812847 | 0 | 30.49508 | 0.0002 | 30.0167 | 0.0002 | 30.30554 | 0.0001 | 0 |
| Libya | LBY | 1036 | 0.0792789 | 0 | 43.98623 | 0 | 43.62646 | 0 | 43.80753 | 0 | 0 |
| Monaco | MCO | 509 | 0.0788709 | 0.0008 | 17.70252 | 0.0243 | 18.34573 | 0.0207 | 17.88676 | 0.0224 | 1 |
| Russian Federation | RUS | 1314 | 0.0787358 | 0 | 76.30465 | 0 | 70.86308 | 0 | 74.263 | 0 | 0 |
| Mali | MLI | 939 | 0.0784512 | 0 | 36.39813 | 0 | 38.01417 | 0 | 36.86747 | 0 | 1 |
| Honduras | HND | 1048 | 0.0772508 | 0 | 35.66253 | 0 | 34.70983 | 0 | 35.29828 | 0 | 0 |
| Côte d'Ivoire | CIV | 892 | 0.0768147 | 0 | 34.227 | 0.0001 | 36.6999 | 0 | 34.97493 | 0.0001 | 0 |
| Zimbabwe | ZWE | 1348 | 0.0759816 | 0 | 43.77814 | 0 | 45.82307 | 0 | 44.38836 | 0 | 0 |
| Pakistan | PAK | 1286 | 0.0750995 | 0 | 40.84404 | 0 | 39.91445 | 0 | 40.50036 | 0 | 0 |
| Trinidad and Tobago | TTO | 895 | 0.0748341 | 0 | 25.53746 | 0.0013 | 26.35955 | 0.0008 | 25.78469 | 0.001 | 0 |
| Singapore | SGP | 848 | 0.0748268 | 0.0001 | 26.28188 | 0.001 | 27.21759 | 0.0005 | 26.56405 | 0.0007 | 0 |
| Uganda | UGA | 927 | 0.0741193 | 0 | 25.61647 | 0.0014 | 26.48057 | 0.0005 | 25.87531 | 0.001 | 0 |
| Zimbabwe | ZWE | 988 | 0.0740534 | 0.0001 | 33.71384 | 0 | 34.19126 | 0 | 33.8249 | 0 | 0 |
| Viet Nam | VNM | 796 | 0.0738194 | 0 | 25.96649 | 0.0015 | 26.79475 | 0.001 | 26.20973 | 0.0012 | 0 |
| Iran | IRN | 1365 | 0.0737945 | 0 | 67.11988 | 0 | 70.90876 | 0 | 68.24692 | 0 | 0 |
| Djibouti | DJI | 594 | 0.0733838 | 0.001 | 16.64701 | 0.0352 | 16.91643 | 0.034 | 16.71926 | 0.0343 | 1 |
| Haiti | HTI | 820 | 0.0711107 | 0 | 27.48284 | 0.0004 | 30.55704 | 0.0002 | 28.41537 | 0.0003 | 1 |
| United Kingdom | GBR | 1364 | 0.0708247 | 0 | 47.94358 | 0 | 51.01892 | 0 | 48.86348 | 0 | 0 |
| Albania | ALB | 1171 | 0.0700361 | 0 | 51.01077 | 0 | 51.89535 | 0 | 51.19775 | 0 | 0 |
| Sweden | SWE | 818 | 0.0700308 | 0.0001 | 40.24113 | 0 | 37.45435 | 0 | 39.22022 | 0 | 0 |
| Kuwait | KWT | 1197 | 0.0698973 | 0 | 43.90362 | 0 | 43.55134 | 0 | 43.72731 | 0 | 0 |
| Benin | BEN | 220 | 0.0692118 | 0.0951 | 9.094197 | 0.3357 | 10.51491 | 0.2412 | 9.512335 | 0.3027 | 0 |
| Oman | OMN | 859 | 0.0691679 | 0.0001 | 36.21967 | 0 | 36.5141 | 0 | 36.25905 | 0 | 0 |
| India | IND | 1321 | 0.069097 | 0 | 60.06876 | 0 | 56.73045 | 0 | 58.86046 | 0 | 0 |
| Mauritania | MRT | 909 | 0.0689068 | 0 | 27.40192 | 0.0009 | 30.40529 | 0.0005 | 28.31434 | 0.0006 | 1 |
| Kosovo | RKS | 901 | 0.0685593 | 0.0004 | 38.09676 | 0.0001 | 38.75871 | 0.0001 | 38.25619 | 0.0001 | 0 |
| Niger | NER | 640 | 0.0681912 | 0.0014 | 19.10217 | 0.0136 | 19.94327 | 0.0114 | 19.34347 | 0.0123 | 1 |
| Belize | BLZ | 614 | 0.068131 | 0.0017 | 16.49737 | 0.0335 | 17.72427 | 0.0219 | 16.87317 | 0.0289 | 0 |
| Netherlands | NLD | 1306 | 0.0660722 | 0 | 41.66957 | 0 | 40.17288 | 0 | 41.12632 | 0 | 0 |
| Greece | GRC | 1243 | 0.06531 | 0 | 47.55806 | 0 | 46.84926 | 0 | 47.26962 | 0 | 0 |
| Namibia | NAM | 947 | 0.0642547 | 0 | 20.51633 | 0.0096 | 21.2838 | 0.0075 | 20.75285 | 0.009 | 0 |
| Andorra | AND | 532 | 0.0641879 | 0.0061 | 14.83716 | 0.0622 | 15.85845 | 0.0445 | 15.14243 | 0.055 | 1 |
| Guinea | GIN | 683 | 0.0635381 | 0.0019 | 15.8969 | 0.043 | 15.7783 | 0.0458 | 15.84464 | 0.0437 | 1 |
| Philippines | PHL | 1324 | 0.0630183 | 0 | 48.15043 | 0 | 50.69274 | 0 | 48.9023 | 0 | 0 |
| Kyrgyzstan | KGZ | 1087 | 0.0627595 | 0.0002 | 24.56171 | 0.0023 | 25.16426 | 0.0017 | 24.74367 | 0.0021 | 0 |
| Slovenia | SVN | 1135 | 0.0620858 | 0 | 30.15661 | 0.0001 | 31.08853 | 0.0001 | 30.42187 | 0 | 0 |
| Iceland | ISL | 515 | 0.0620768 | 0.0088 | 18.65353 | 0.0145 | 19.07766 | 0.0127 | 18.77652 | 0.014 | 0 |
| France | FRA | 1300 | 0.0617992 | 0.0001 | 47.31948 | 0 | 49.66996 | 0 | 47.97357 | 0 | 0 |
| Saint Vincent and the Grenadines | VCT | 320 | 0.06147 | 0.0637 | 12.89166 | 0.1143 | 14.42395 | 0.0731 | 13.32988 | 0.1011 | 1 |
| Lesotho | LSO | 377 | 0.06107 | 0.0434 | 15.5756 | 0.0535 | 16.90414 | 0.0352 | 15.97436 | 0.0448 | 0 |
| Austria | AUT | 1303 | 0.0594903 | 0 | 33.42154 | 0.0001 | 33.20553 | 0.0001 | 33.30856 | 0.0001 | 0 |
| Angola | AGO | 1057 | 0.0592456 | 0.0001 | 25.31863 | 0.0011 | 26.41475 | 0.0008 | 25.6412 | 0.0008 | 0 |
| Burundi | BDI | 377 | 0.0586878 | 0.0527 | 24.1749 | 0.0016 | 22.26292 | 0.0041 | 23.46371 | 0.002 | 0 |
| Venezuela | VEN | 1144 | 0.0583537 | 0.0004 | 76.77615 | 0 | 84.79006 | 0 | 79.11019 | 0 | 0 |
| Dominican Republic | DOM | 1213 | 0.0583249 | 0.0001 | 25.18226 | 0.001 | 24.40072 | 0.0016 | 24.89282 | 0.0013 | 0 |
| Argentina | ARG | 1313 | 0.0575322 | 0.0001 | 25.41614 | 0.0016 | 25.54894 | 0.0012 | 25.44617 | 0.0014 | 0 |
| El Salvador | SLV | 1042 | 0.0559947 | 0.0006 | 50.84316 | 0 | 48.91 | 0 | 50.06728 | 0 | 0 |
| Senegal | SEN | 1085 | 0.0547304 | 0.0005 | 17.78331 | 0.0208 | 17.54012 | 0.0224 | 17.69312 | 0.0214 | 0 |
| South Africa | ZAF | 1293 | 0.0539584 | 0.0004 | 32.60761 | 0.0001 | 32.76339 | 0.0002 | 32.63283 | 0.0001 | 0 |
| North Macedonia | MKD | 1257 | 0.0536155 | 0.0004 | 30.91735 | 0.0002 | 31.48634 | 0.0002 | 31.08487 | 0.0002 | 0 |
| Armenia | ARM | 1287 | 0.0535703 | 0.0002 | 28.02299 | 0.0007 | 30.05927 | 0.0005 | 28.64772 | 0.0007 | 0 |
| Eswatini | SWZ | 892 | 0.0531478 | 0.0034 | 14.3201 | 0.0741 | 14.82649 | 0.0629 | 14.47842 | 0.0694 | 0 |
| Colombia | COL | 1305 | 0.0531124 | 0.0002 | 22.32726 | 0.0037 | 22.95692 | 0.0026 | 22.52053 | 0.0033 | 0 |
| Uruguay | URY | 1068 | 0.0528413 | 0.0014 | 34.67709 | 0 | 35.89613 | 0 | 35.03749 | 0 | 0 |
| Zambia | ZMB | 1240 | 0.0527175 | 0 | 20.36938 | 0.0084 | 20.75462 | 0.007 | 20.48425 | 0.0077 | 0 |
| Estonia | EST | 1032 | 0.0527027 | 0.0017 | 19.76161 | 0.0119 | 19.48896 | 0.0124 | 19.65578 | 0.0119 | 0 |

| | | | | | | | | | | | |
|---|---|---|---|---|---|---|---|---|---|---|---|
| New Zealand | NZL | 538 | 0.0526185 | 0.0348 | 11.1347 | 0.1964 | 11.70383 | 0.1691 | 11.31261 | 0.1867 | 1 |
| Sri Lanka | LKA | 1059 | 0.0524849 | 0.0015 | 55.52604 | 0 | 53.64893 | 0 | 54.78587 | 0 | 0 |
| Rwanda | RWA | 971 | 0.0522141 | 0.0021 | 20.59321 | 0.0085 | 20.79623 | 0.0076 | 20.64866 | 0.0077 | 0 |
| Guyana | GUY | 944 | 0.0520893 | 0.0025 | 18.95891 | 0.0145 | 19.2153 | 0.0132 | 19.02676 | 0.0141 | 1 |
| Sudan | SDN | 601 | 0.0519969 | 0.0265 | 10.13065 | 0.2544 | 10.39452 | 0.2421 | 10.21203 | 0.2504 | 0 |
| Botswana | BWA | 295 | 0.0516923 | 0.1836 | 5.022262 | 0.7525 | 5.230846 | 0.7329 | 5.087487 | 0.7457 | 0 |
| Canada | CAN | 1346 | 0.0514155 | 0.0004 | 30.94407 | 0.0001 | 30.42519 | 0.0002 | 30.7469 | 0.0002 | 0 |
| Hungary | HUN | 1134 | 0.04994 | 0.002 | 40.10039 | 0 | 40.13517 | 0 | 40.05602 | 0 | 0 |
| Panama | PAN | 1255 | 0.0498688 | 0.0009 | 20.97067 | 0.0077 | 20.25296 | 0.0101 | 20.7151 | 0.0086 | 0 |
| Comoros | COM | 394 | 0.049761 | 0.1124 | 11.50325 | 0.1711 | 12.01255 | 0.1527 | 11.64324 | 0.1648 | 1 |
| Ireland | IRL | 1036 | 0.0495151 | 0.0023 | 36.17774 | 0 | 35.09364 | 0 | 35.77814 | 0 | 0 |
| Ghana | GHA | 664 | 0.0493491 | 0.0266 | 10.27061 | 0.244 | 10.05421 | 0.2618 | 10.19274 | 0.2492 | 0 |
| Malawi | MWI | 900 | 0.04897 | 0.0059 | 18.2953 | 0.0188 | 19.19279 | 0.0145 | 18.56735 | 0.0174 | 0 |
| Bahrain | BHR | 1025 | 0.0485605 | 0.0041 | 27.07416 | 0.0014 | 30.6141 | 0 | 28.12843 | 0.0007 | 0 |
| Lebanon | LBN | 1211 | 0.0482681 | 0.0008 | 37.03782 | 0.0001 | 37.43488 | 0.0001 | 37.12387 | 0.0001 | 0 |
| Indonesia | IDN | 1320 | 0.0482124 | 0.0013 | 34.35292 | 0 | 35.47361 | 0.0001 | 34.69497 | 0 | 0 |
| Yemen | YEM | 909 | 0.0480047 | 0.0088 | 12.08011 | 0.148 | 12.64863 | 0.1245 | 12.25893 | 0.1392 | 1 |
| Kazakhstan | KAZ | 825 | 0.0475223 | 0.0147 | 27.8306 | 0.0005 | 26.91261 | 0.0009 | 27.45664 | 0.0006 | 0 |
| Thailand | THA | 987 | 0.0473828 | 0.0071 | 18.44533 | 0.0166 | 19.90921 | 0.0087 | 18.89971 | 0.0138 | 0 |
| China | CHN | 922 | 0.0472284 | 0.0098 | 45.05334 | 0 | 43.4539 | 0 | 44.4184 | 0 | 0 |
| Jamaica | JAM | 1077 | 0.0472234 | 0.0047 | 18.63587 | 0.0168 | 17.55835 | 0.0256 | 18.25288 | 0.0195 | 0 |
| Kosovo | RKS | 968 | 0.0471105 | 0.0062 | 14.14682 | 0.0801 | 14.2631 | 0.0795 | 14.17722 | 0.0794 | 0 |
| Congo, Democratic Republic | COD | 795 | 0.0469966 | 0.0176 | 15.67336 | 0.0484 | 15.67626 | 0.0477 | 15.65833 | 0.0478 | 0 |
| Suriname | SUR | 895 | 0.0469011 | 0.0118 | 18.76694 | 0.0155 | 19.35098 | 0.0124 | 18.94834 | 0.0145 | 0 |
| Cabo Verde | CPV | 851 | 0.0467961 | 0.0146 | 25.23047 | 0.0016 | 25.04249 | 0.0021 | 25.14481 | 0.0016 | 1 |
| Slovakia | SVK | 1061 | 0.0467551 | 0.0057 | 16.79992 | 0.0325 | 16.15684 | 0.0396 | 16.57459 | 0.0338 | 0 |
| Kenya | KEN | 1216 | 0.0451871 | 0.0037 | 13.56673 | 0.091 | 13.3688 | 0.0985 | 13.49657 | 0.0935 | 0 |
| Moldova, Republic | MDA | 1295 | 0.0441437 | 0.0034 | 14.01416 | 0.0813 | 13.86424 | 0.086 | 13.95949 | 0.0821 | 0 |
| Algeria | DZA | 1321 | 0.0434326 | 0.0036 | 37.83593 | 0 | 35.34669 | 0 | 36.93475 | 0 | 0 |
| Morocco | MAR | 1278 | 0.042849 | 0.0048 | 24.78115 | 0.0021 | 24.35545 | 0.0027 | 24.6244 | 0.0022 | 0 |
| Ethiopia | ETH | 1210 | 0.0427717 | 0.0073 | 31.18589 | 0.0001 | 32.24725 | 0.0001 | 31.50853 | 0.0001 | 0 |
| Nigeria | NGA | 1079 | 0.0418801 | 0.0135 | 32.85805 | 0 | 33.90879 | 0 | 33.1653 | 0 | 0 |
| Belgium | BEL | 1181 | 0.0414931 | 0.0093 | 18.72678 | 0.0176 | 19.14446 | 0.0156 | 18.85243 | 0.0166 | 0 |
| Ukraine | UKR | 1305 | 0.0408864 | 0.0077 | 20.81486 | 0.0085 | 21.74421 | 0.0066 | 21.10469 | 0.0073 | 0 |
| Jordan | JOR | 1092 | 0.0407047 | 0.0174 | 23.96128 | 0.0033 | 23.98903 | 0.0028 | 23.94923 | 0.0034 | 0 |
| Korea, Republic | KOR | 1295 | 0.0406162 | 0.0086 | 43.98446 | 0 | 46.70509 | 0 | 44.75086 | 0 | 0 |
| Cambodia | KHM | 707 | 0.0405324 | 0.0737 | 27.29135 | 0.001 | 26.0811 | 0.0014 | 26.83359 | 0.001 | 0 |
| Guatemala | GTM | 1258 | 0.0402987 | 0.0106 | 21.50378 | 0.0077 | 21.16923 | 0.0084 | 21.38101 | 0.0079 | 0 |
| Palestine | PSE | 1061 | 0.0388448 | 0.0266 | 10.62131 | 0.2198 | 11.01948 | 0.1984 | 10.74768 | 0.2128 | 0 |
| Portugal | PRT | 1308 | 0.0387976 | 0.0106 | 17.06297 | 0.029 | 16.76493 | 0.0336 | 16.95306 | 0.0303 | 0 |
| Serbia | SRB | 1277 | 0.0381786 | 0.0136 | 51.30633 | 0 | 52.62306 | 0 | 51.62467 | 0 | 0 |
| Norway | NOR | 913 | 0.0377059 | 0.0553 | 18.61418 | 0.0185 | 20.08314 | 0.0106 | 19.04487 | 0.0155 | 0 |
| Brazil | BRA | 1324 | 0.0375442 | 0.0114 | 17.43218 | 0.024 | 17.16064 | 0.0261 | 17.33062 | 0.0246 | 0 |
| Ecuador | ECU | 1248 | 0.0347523 | 0.0307 | 16.7342 | 0.0319 | 15.77222 | 0.0452 | 16.38825 | 0.036 | 0 |
| Papua New Guinea | PNG | 378 | 0.0338342 | 0.4401 | 7.855677 | 0.4496 | 8.751019 | 0.3655 | 8.124021 | 0.423 | 1 |
| Bulgaria | BGR | 1275 | 0.0327947 | 0.0436 | 13.41701 | 0.0973 | 13.80124 | 0.0869 | 13.53763 | 0.0933 | 0 |
| Georgia | GEO | 1148 | 0.0319993 | 0.0676 | 25.6556 | 0.0017 | 24.84625 | 0.002 | 25.36232 | 0.0019 | 0 |
| Seychelles | SYC | 376 | 0.0314168 | 0.5012 | 4.857517 | 0.7705 | 4.977145 | 0.7586 | 4.894531 | 0.7669 | 1 |
| Romania | ROU | 1318 | 0.0312917 | 0.0531 | 13.81752 | 0.084 | 14.06807 | 0.0783 | 13.89642 | 0.0829 | 0 |
| Somalia | SOM | 471 | 0.0303097 | 0.4287 | 11.23996 | 0.1889 | 10.97573 | 0.2045 | 11.13367 | 0.1945 | 1 |
| Croatia | HRV | 1225 | 0.030004 | 0.0902 | 20.79246 | 0.0082 | 21.40227 | 0.0063 | 20.9704 | 0.0072 | 0 |
| Cameroon | CMR | 304 | 0.029902 | 0.625 | 8.659315 | 0.3657 | 9.813681 | 0.281 | 8.992525 | 0.3385 | 0 |
| Latvia | LVA | 1077 | 0.0297761 | 0.1229 | 16.48287 | 0.0365 | 16.55351 | 0.0353 | 16.49579 | 0.0358 | 0 |
| Switzerland | CHE | 1086 | 0.0295409 | 0.1178 | 10.131 | 0.2558 | 10.17035 | 0.2563 | 10.1414 | 0.2556 | 0 |
| Italy | ITA | 1364 | 0.0295354 | 0.0682 | 27.21724 | 0.0008 | 28.21639 | 0.0004 | 27.51437 | 0.0007 | 0 |
| Chile | CHL | 1311 | 0.0292148 | 0.0826 | 9.740234 | 0.2808 | 9.646721 | 0.2863 | 9.705089 | 0.2822 | 0 |
| Tunisia | TUN | 1085 | 0.0280023 | 0.1546 | 10.84227 | 0.2114 | 10.98003 | 0.2054 | 10.88396 | 0.2103 | 0 |
| Finland | FIN | 1118 | 0.0262481 | 0.1843 | 16.27464 | 0.036 | 17.04577 | 0.0274 | 16.50541 | 0.0324 | 0 |
| Madagascar | MDG | 636 | 0.0259711 | 0.4362 | 9.408001 | 0.3092 | 9.696239 | 0.2931 | 9.489809 | 0.304 | 0 |
| Japan | JPN | 1336 | 0.0249642 | 0.1546 | 16.67792 | 0.03 | 16.56973 | 0.0327 | 16.63025 | 0.031 | 0 |
| Poland | POL | 1298 | 0.0221394 | 0.2609 | 18.51897 | 0.0181 | 19.76265 | 0.0117 | 18.89911 | 0.0157 | 0 |
| Bolivia | BOL | 1198 | 0.0211737 | 0.3297 | 18.50615 | 0.0185 | 18.21397 | 0.0206 | 18.39194 | 0.019 | 0 |
| Germany | DEU | 1339 | 0.0202649 | 0.3134 | 8.874732 | 0.3493 | 9.145521 | 0.3262 | 8.959892 | 0.3421 | 0 |
| Spain | ESP | 986 | 0.0199099 | 0.4797 | 6.131133 | 0.6309 | 5.954826 | 0.6535 | 6.070333 | 0.6385 | 0 |
| Afghanistan | AFG | 1148 | 0.0193839 | 0.4376 | 8.365998 | 0.387 | 8.55317 | 0.3709 | 8.421782 | 0.3816 | 0 |
| Malaysia | MYS | 1211 | 0.0186509 | 0.4392 | 9.187576 | 0.3215 | 9.274173 | 0.3142 | 9.212535 | 0.3184 | 0 |

| | | | | | | | | | | | |
|---|---|---|---|---|---|---|---|---|---|---|---|
| United States of America | USA | 1356 | 0.0177165 | 0.4372 | 3.976688 | 0.859 | 4.044902 | 0.8533 | 3.99886 | 0.8562 | 0 |
| Bosnia and Herzegovina | BIH | 1017 | 0.0168609 | 0.6238 | 6.367772 | 0.6021 | 6.21468 | 0.6216 | 6.314681 | 0.6087 | 0 |
| Denmark | DNK | 1140 | 0.0161507 | 0.6003 | 13.16769 | 0.1097 | 13.29318 | 0.1057 | 13.20231 | 0.1086 | 0 |
| Israel | ISR | 1232 | 0.01329 | 0.7279 | 16.01816 | 0.0422 | 16.1672 | 0.0409 | 16.05158 | 0.0414 | 0 |

#### 8. Measuring information-gathering capacity

I constructed an indicator of each country's ability to collect and process data based on the latent factor analysis of three input variables:

(1) Hanson and Sigman's (2021) measure of census frequency captures the number of years between each national census.
(2) The World Bank's (2022) Statistical Capacity index is a composite score capturing methodology, data sources, as well as periodicity and timeliness.
(3) Brambor and colleague's (2020) information capacity index is based on whether a country has a national statistical agency, a civil register, a population register, as well as its ability to produce a census and statistical yearbook.

All three inputs are for the year 2015. The latent factor analysis was carried out using structural equation modelling, with full information maximum likelihood. The factor loadings were checked to ensure they are statistically significant and of sufficient magnitude. In addition, goodness of fit statistics were used to check whether the latent variable was sufficiently related to the input variables. This procedure allowed me to construct an overall index for 174 countries.

#### References

Bjørnskov, Christian, and Martin Rode. 2020. “Regime Types and Regime Change: A New Dataset on Democracy, Coups, and Political Institutions.” *The Review of International Organizations* 15 (2): 531–51. https://doi.org/10.1007/s11558-019-09345-1.

Cheibub, José Antonio, Jennifer Gandhi, and James Raymond Vreeland. 2009. “Democracy and Dictatorship Revisited.” *Public Choice* 143 (1–2): 67–101. https://doi.org/10.1007/s11127-009-9491-2.

Coppedge, Michael, and et al. 2023. “V-Dem [Country-Year/Country-Date] Dataset V13.” Data set. Varieties of Democracy (V-Dem) Project. University of Gothenburg: Varieties of Democracy Institute. https://doi.org/10.23696/vdemds23.

Diekmann, Andreas. 2007. “Not the First Digit! Using Benford’s Law to Detect Fraudulent Scientific Data.” *Journal of Applied Statistics* 34 (3): 321–29. https://doi.org/10.1080/02664760601004940.

Dong, Ensheng, Hongru Du, and Lauren Gardner. 2020. “An Interactive Web-Based Dashboard to Track COVID-19 in Real Time.” *The Lancet Infectious Diseases* 20 (5): 533–34. https://doi.org/10.1016/S1473-3099(20)30120-1.

Fewster, R. M. 2009. “A Simple Explanation of Benford’s Law.” *The American Statistician* 63 (1): 26–32. https://doi.org/10.1198/tast.2009.0005.

GBD Collaborative Network. 2020a. “Global Burden of Disease Study 2019 (GBD 2019) Covariates 1980-2019.” Data set. Global Burden of Disease. Seattle, United States: Institute for Health Metrics and Evaluation (IHME), University of Washington. https://doi.org/10.6069/CFCY-WA51.

———. 2020b. “Global Burden of Disease Study 2019 (GBD 2019) Results.” Data set. Seattle, United States: Institute for Health Metrics and Evaluation (IHME), University of Washington. http://ghdx.healthdata.org/gbd-results-tool.

Goodman, William. 2016. “The Promises and Pitfalls of Benford’s Law.” *Significance* 13 (3): 38–41. https://doi.org/10.1111/j.1740-9713.2016.00919.x.

Gründler, Klaus, and Tommy Krieger. 2021. “Using Machine Learning for Measuring Democracy: A Practitioners Guide and a New Updated Dataset for 186 Countries from 1919 to 2019.” *European Journal of Political Economy* 70 (December): 102047. https://doi.org/10.1016/j.ejpoleco.2021.102047.

Guha-Sapir, Debarati. 2022. “EM-DAT. The Emergency Events Database.” Data set. Brussels, Belgium: Université Catholique de Louvain (UCL) - CRED. https://www.emdat.be/.

Heritage Foundation. n.d. “Economic Data and Statistics on World Economy and Economic Freedom.” Dataset. Heritage Foundation. Accessed March 26, 2021. https://www.heritage.org/index/download.

Karlinsky, Ariel, and Dmitry Kobak. 2021. “Tracking Excess Mortality across Countries during the COVID-19 Pandemic with the World Mortality Dataset.” *ELife* 10 (June): e69336. https://doi.org/10.7554/eLife.69336.

Kobak, Dmitry. 2022. “Underdispersion: A Statistical Anomaly in Reported Covid Data.” *Significance* 19 (2): 10–13. https://doi.org/10.1111/1740-9713.01627.

Marshall, Monty G., and Ted Robert Gurr. 2020. “Polity5: Political Regime Characteristics and Transitions, 1800-2018.” Data set Polity5 dataset version 2018. Center for Systemic Peace. http://www.systemicpeace.org/inscrdata.html.

Michalski, Tomasz, and Gilles Stoltz. 2013. “Do Countries Falsify Economic Data Strategically? Some Evidence That They Might.” *Review of Economics and Statistics* 95 (2): 591–616. https://doi.org/10.1162/REST_a_00274.

Pettersson, Therése, Shawn Davies, Amber Deniz, Garoun Engström, Nanar Hawach, Stina Högbladh, and Margareta Sollenberg Magnus Öberg. 2021. “Organized Violence 1989–2020, with a Special Emphasis on Syria.” *Journal of Peace Research* 58 (4): 809–25. https://doi.org/10.1177/00223433211026126.

Skaaning, Svend-Erik, John Gerring, and Henrikas Bartusevičius. 2015. “A Lexical Index of Electoral Democracy.” *Comparative Political Studies* 48 (12): 1491–1525. https://doi.org/10.1177/0010414015581050.

The Economist. (2020) 2022. “The Economist’s Tracker for Covid-19 Excess Deaths.” Data set & code. The Economist. https://github.com/TheEconomist/covid-19-excess-deaths-tracker.

UNDESA. 2022. “World Population Prospects 2022.” Data set. New York, NY: United Nations Department of Economic and Social Affairs. https://population.un.org/wpp/.

Wang, Haidong, Katherine R. Paulson, Spencer A. Pease, Stefanie Watson, Haley Comfort, Peng Zheng, Aleksandr Y. Aravkin, et al. 2022. “Estimating Excess Mortality Due to the COVID-19 Pandemic: A Systematic Analysis of COVID-19-Related Mortality, 2020–21.” *The Lancet* 399 (10334): 1513–36. https://doi.org/10.1016/S0140-6736(21)02796-3.

WHO. 2021. “Index of Service Capacity and Access.” Data set. Global Health Observatory. Geneva: World Health Organization. https://www.who.int/data/gho/data/indicators/indicator-details/GHO/uhc-sci-components-service-capacity-and-access.

———. 2022. “Global Excess Deaths Associated with COVID-19 (Modelled Estimates).” Data set. Geneva: World Health Organization. https://www.who.int/data/sets/global-excess-deaths-associated-with-covid-19-modelled-estimates.

World Bank. 2022. “Statistical Capacity Indicator.” Data set. Washington D.C.: World Bank. https://datatopics.worldbank.org/statisticalcapacity/.

———. n.d. “The World by Income and Region.” World Development Indicators. Accessed October 6, 2021. https://datatopics.worldbank.org/world-development-indicators/the-world-by-income-and-region.html.
